# Cortical signatures of sleep are altered following effective deep brain stimulation for depression

**DOI:** 10.1101/2023.12.08.23299724

**Authors:** Joram J. van Rheede, Sankaraleengam Alagapan, Timothy J. Denison, Patricio Riva-Posse, Christopher J. Rozell, Helen S. Mayberg, Allison C. Waters, Andrew Sharott

## Abstract

Deep brain stimulation (DBS) of the subcallosal cingulate cortex (SCC) is an experimental therapy for treatment-resistant depression (TRD). Chronic SCC DBS leads to long-term changes in the electrophysiological dynamics measured from local field potential (LFP) during wakefulness, but it is unclear how it impacts sleep-related brain activity. This is a crucial gap in knowledge, given the link between depression and sleep disturbances, and an emerging interest in the interaction between DBS, sleep, and circadian rhythms. We therefore sought to characterise changes in electrophysiological markers of sleep associated with DBS treatment for depression. We analysed key electrophysiological signatures of sleep – slow-wave activity (SWA, 0.5-4.5Hz) and sleep spindles – in LFPs recorded from the SCC of 9 patients who responded to DBS for TRD. This allowed us to compare the electrophysiological changes before and after 24 weeks of therapeutically effective SCC DBS.

SWA power was highly correlated between hemispheres, consistent with a global sleep state. Furthermore, SWA occurred earlier in the night after chronic DBS and had a more prominent peak. While we found no evidence for changes to slow wave power or stability, we found an increase in the density of sleep spindles. Our results represent a first-of-its-kind report on long-term electrophysiological markers of sleep recorded from the SCC in patients with TRD, and provides evidence of earlier NREM sleep and increased sleep spindle activity following clinically effective DBS treatment. Future work is needed to establish the causal relationship between long-term DBS and the neural mechanisms underlying sleep.

**Conflict of interest statement:** JJvR has received honoraria from Medtronic. TD is a founder and director of Amber who own Bioinduction Ltd, has received honoraria from Medtronic, and is an advisor for Cortec Neuro and Synchron. PR-P receives consulting fees from Abbott Labs, LivaNova, and Janssen Pharmaceuticals. HM receives consulting and Intellectual Property licensing fees from Abbott Labs. SA, CR, ACW and AS have nothing to declare.

## Introduction

Deep brain stimulation (DBS) of the subcallosal cingulate cortex (SCC) is a promising neuromodulation therapy for treatment-resistant depression (TRD), with open-label studies repeatedly reporting efficacy [1–5]. SCC DBS has been shown to improve the constellation of symptoms underlying TRD, including depressed mood, anxiety, and sleep [1,2]. However, it is not clear if SCC DBS affects the electrophysiological characteristics of sleep. This is an important gap in knowledge as disruption in sleep is an established symptom of major depressive disorder (MDD)[6]. The relationship between MDD and sleep disturbances is bidirectional [7–14], and sleep disorders predict poorer treatment outcomes and likelihood of relapse [15–18]. There is also evidence that sleep *structure* is altered in depression, with reduced occurrence of the deeper stages of NREM and therefore slow-wave [19–24] activity especially during the earlier sleep cycles, and a decreased REM sleep latency. These sleep EEG properties appear to track depressive symptomatology [25–27], though there is some evidence this is moderated by age and gender [28,29]. Moreover, even without self-reported sleep problems, an evening-oriented circadian preference (late sleeping and late waking) is associated with an increased risk for depression and worse symptoms [15,30–33], and there is increasing evidence that circadian disruption and neuropsychiatric conditions have common mechanisms [34]. Given this strong link between depression and sleep quality, structure and pattern, it is important to understand if SCC DBS alters patients’ sleep or sleep-wake cycle. Insights into this interaction will help improve therapeutic decisions such as the selection of optimal stimulation parameters for sleep [35,36], and could inform the development of adaptive controllers that adjust stimulation parameters according to endogenous biorhythms [37].

Advances in neurotechnology have enabled implanted DBS pulse generators that can record local field potentials (LFP) from the stimulation target region while patients go about their daily lives [38,39]. In addition to providing an opportunity to investigate the electrophysiological changes associated with DBS during wakefulness [40–42], these recordings allow for the investigation of changes in electrophysiological signatures of sleep and fluctuations related to the time of day [43]. The sleep-wake cycle is the most prominent source of intra-day variability in human cortical activity [44], but has not yet been characterised using chronic recordings on an implanted DBS device. Non-rapid-eye-movement (NREM) sleep in particular is characterised by cortical activity not normally observed during waking, including prominent high amplitude slow wave activity [44,45] and sleep spindles [46–50]. While polysomnography remains the gold standard evaluation of sleep quality, increased understanding of sleep signatures from invasive recordings [51] allows inference of broad changes in sleep and sleep-wake cycle over the course of treatment with DBS.

We examined LFP data from the SCC acquired from an implanted DBS device to investigate the effects of SCC DBS on sleep and to identify potential sleep-related biomarkers of treatment response [FDA IDE G130107, Clinical Trials. NCT01984710]. LFP data were recorded longitudinally from a cohort of 10 patients with TRD as they underwent chronic SCC DBS. The high response rate (90%) at the primary endpoint in this cohort allows us to evaluate changes that accompany effective DBS therapy [42,52]. Clinical response was defined as a 50% reduction in HDRS vs baseline. In a previous report, we showed how changes in SCC dynamics tracked recovery in this cohort [42]. Here we report changes in sleep signatures and the sleep-wake cycle measured from LFP in the 9 treatment responders after 24 weeks of therapeutic DBS.

## Materials and methods

### Patient and trial details

Data reported here were collected as part of an experimental trial to derive biomarkers to optimize SCC DBS for TRD (ClinicalTrials.gov: NCT01984710). Ten subjects with treatment-resistant major depressive disorder were consecutively enrolled. All participants provided written informed consent, and the protocol adhered to the Declaration of Helsinki. The protocol was approved by the Institutional Review Boards of Emory University, Icahn School of Medicine at Mount Sinai and Georgia Institute of Technology, as well as the US Food and Drug Administration under a physician-sponsored Investigational Device Exemption (IDE G130107), with monitoring by the Emory University Department of Psychiatry and Behavioral Sciences Data and Safety Monitoring Board.

The study design included the following phases: 1) four weeks of presurgical baseline, during which only clinical assessments were performed, 2) at least four weeks of recovery following surgical implantation of the device, during which LFP was recorded daily with stimulation turned OFF, 3) twenty-four weeks of therapeutic DBS, and 4) one week when stimulation was discontinued in a single-blind manner (i.e. patient unaware) with daily LFP recordings (Figure 1A). All patients showed improvement on the Hamilton Depression Rating Scale (HDRS-17) following treatment (Table 1), though some improvement was already observed in the Pre-DBS post-operative period, consistent with placebo or lesion effects [53].

**Figure 1:**
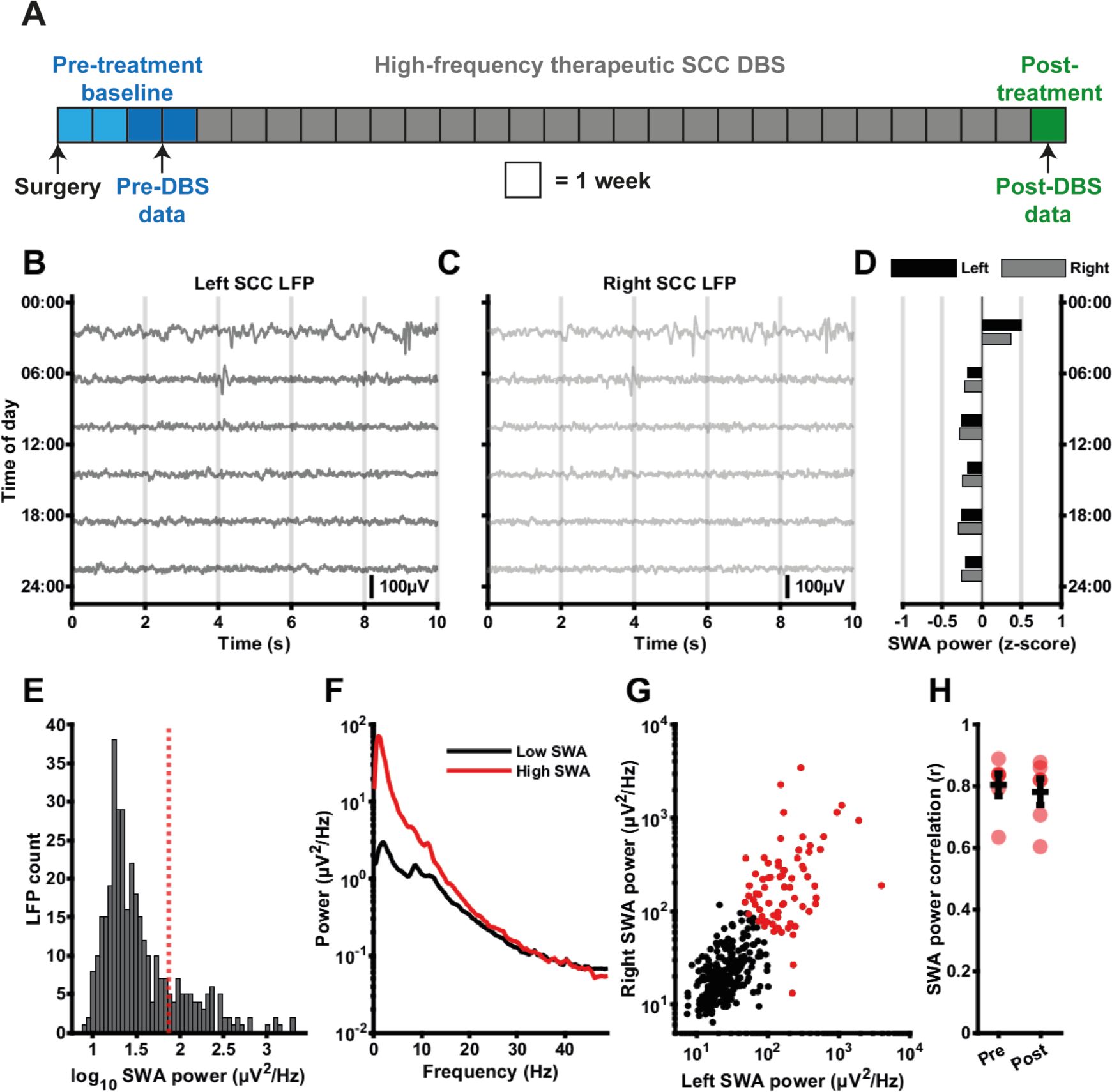
Quantifying slow-wave activity in bilateral SCC LFP segments collected at different times of day. **A:** Overview of the data collection timeline. LFP segments from weeks 3 and 4 after surgery, but before initiation of therapeutic DBS comprised the Pre-DBS baseline phase (mean number of Pre-DBS samples 121±59; typical sampling frequency 1 sample / 3.06 hours), while LFP segments recorded 1 week after 24 weeks of therapeutic DBS (during which DBS was switched off) were used for the Post-DBS phase (mean number of Post-DBS samples 386±74; typical sampling frequency 1 sample / 0.375 hours). **B, C:** Left (B) and right (C) SCC LFP segments (low-pass filtered with a cut-off at 30Hz) collected at different times of a single day from one example patient during the Pre-DBS phase. Scale bar represents 100uV. **D:** SWA (0.5-4.5Hz) power z-scores corresponding to the left (black) and right (grey) hemisphere LFP segments shown in B&C. **E:** Histogram of log_10_ SWA power (averaged between left and right) for all Pre-DBS LFP segments for one example patient. The red dotted line represents the 80^th^ percentile that was used as a cut-off for ‘high SWA’ putative sleep episodes. **F:** Average power spectrum (Welch’s method) of ‘high SWA’ (>80^th^ percentile) LFP segments compared to ‘low SWA’ (<50^th^ percentile) LFP segments. **G:** Scatter plot of the correlation between left and right hemisphere SWA power. Red data points represent ‘high SWA’ LFPs. **H:** Correlation coefficients of left and right SWA power for all 6 patients for which bilateral data was obtained, for the Pre-DBS and Post-DBS phase (Individual data points in red, mean and standard deviation provided in black; Pre-DBS: r = 0.805±0.0889, Post-DBS: r = 0.782±0.105, signed-rank = 17, p = 0.219, n = 6).

**Table 1:** Patient demographic information and primary clinical measures. Baseline vs Pre-DBS: t(8) = 6.81, p = 0.00014 Pre-DBS vs. Week 24: t(8) = 7.87, p = 4.9*10^-5^ Baseline vs Week 24: t(8) = 12.9, p = 1.25*10^-6^

**Baseline vs Week 24: $t(8) = 12.9$ , $p = 1.25 \times 10^{-6}$**
| Participant ID | Age at time of surgery (years) | Sex | Pre-surgical baseline HDRS-17 (mean across 4 weeks)<br>Mean: 22.2±1.8 | HDRS-17 Pre-DBS; Weeks 3&4 post-surgery<br>Mean: 17.6±2.2 | HDRS-17 at week 24 of treatment<br>Mean: 6.44±2.55 | Hemispheres included in further analysis |
| --- | --- | --- | --- | --- | --- | --- |
| P001 | 41-45 | F | 23.5 | 20.5 | 3 | Left |
| P002 | 41-45 | F | 20.5 | 14.5 | 8 | Left & Right |
| P003 | 56-60 | M | 19.25 | 15.5 | 6 | Right |
| P004 | 56-60 | F | 23.5 | 19.5 | 7 | Left & Right |
| P005 | 51-55 | M | 20.5 | 18.0 | 10 | - |
| P006 | 56-60 | F | 23.25 | 14.5 | 10 | Left & Right |
| P007 | 36-40 | M | 22.75 | 18.0 | 4 | Left & Right |
| P008 | 41-45 | F | 24.75 | 18.5 | 4 | Left & Right |
| P009 | 26-30 | M | 21.75 | 19.0 | 6 | Left & Right |

### Clinical scoring

Clinical symptom severity was evaluated by an independent rater using the 17-item Hamilton Depression Rating Scale (HDRS-17), Montgomery-Åsberg Depression Rating Scale (MADRS), and self-reported Beck Depression Inventory (BDI) during weekly visits to the laboratory. In addition to DBS, participants patients were prescribed with different medications depending on their clinical history, which included antidepressants and benzodiazepines (Supplementary Table 1). The dosage of these medications was held constant throughout the study.

### Surgical implantation, device details and DBS parameters

Participants underwent bilateral implantation of DBS electrodes (3387, Medtronic, Minneapolis, MN) at the intersection of four white matter pathways (forceps minor, cingulum bundle, uncinate fasciculus, and frontostriatal fibers) as described previously [52,54]. Stimulation was delivered using a voltage-controlled implanted pulse generator (Activa PC+S, Medtronic Inc., Minneapolis, MN) that allowed the collection of local field potential from contacts immediately adjacent of the stimulation contact on either side in a differential configuration. DBS therapy started four weeks after the implantation surgery to allow stable recordings of the depressed baseline, as per our past published protocols [3,4]. Therapy consisted of bilateral monopolar stimulation on a single contact per hemisphere at 130 Hz with 90 µs pulse width. Stimulation amplitude, initialized at 3.5 V, was varied by study psychiatrists based on clinical judgement throughout the 24 weeks. Final stimulation dose ranged from 3.5 – 5 V; without change in contact, pulse width or frequency.

### Acquisition and analysis of LFP data

LFPs were acquired in 12 s segments by the Activa PC+S at 422 Hz at regular intervals. In the post-operative phase, ‘Pre-DBS’ LFPs were acquired approximately every 2-4 hours. Data were downloaded from the device weekly. In the Post-DBS phase, LFPs were collected approximately every 20-30 minutes with daily downloads; the precise sampling regime for individual patients was determined pragmatically by the data storage capacity of the device and the interval between successive clinical visits (For each patient’s exact sampling times, see Supplementary Figures S2-S10, panels A&B). We report mean typical sampling frequencies defined as follows: For each patient, we took the median across days of the median sampling interval for each individual day; typical sampling frequency for the study phase was then calculated as the mean across patients. The Pre-DBS typical sampling frequency was 1 LFP / 3.06 hours, while the Post-DBS typical sampling frequency was 1 LFP / 0.375 hours. During clinical visits, the LFP data were downloaded from the device and saved in text format. LFP data were compiled across days and stored in shareable formats (HDF-5, *.mat*).

We obtained SCC LFP data from 18 hemispheres (9 patients). Data from one patient was contaminated by amplifier artifact bilaterally (P005). In P001 and P003, data from only one hemisphere was of adequate quality as the other hemispheres had artifacts driven by electrocardiographic activity. The remaining dataset represented 14 hemispheres (8 patients; See Table 1 for the included hemispheres for each participant). Baseline Pre-DBS recordings were collected between 14 and 28 days post-operatively, prior to DBS therapy (Figure 1A); for one patient (P008) data from only 4 days at the start of this period were available due to operational error. Post-DBS data were obtained after 24 weeks of therapeutic DBS (Figure 1A).

The first 2s of each LFP segment were rejected to remove the amplifier switching artifact. LFP segments with a voltage range that exceeded a Z-score of 6 (calculated across each individual patient’s LFP segments for the Pre-DBS and Post-DBS periods separately) were excluded from further analysis. This resulted in a mean of 121±59 useable LFP segments (10s each) per hemisphere in the Pre-DBS phase and a mean of 386±74 useable LFP segments in the Post-DBS phase.

### Calculation of spectral power

SWA power was calculated as band power in the 0.5-4.5Hz range for each LFP segment and each hemisphere using the MATLAB *bandpower()* function. Spindle band power (for Figure 3) was similarly calculated as power in the 9-16Hz range. To determine the peak frequency of spindle-like activity during LFPs collected during putative NREM sleep, we used continuous wavelet transform (MATLAB’s *cwt()*) to calculate a global wavelet spectrum by taking the square of the absolute value of the wavelet transform and taking the mean across the time dimension.

### Stability of SWA

The stability of SWA was quantified using the Temporal Variability Index (TVI), a measure of oscillatory stability independent of overall signal power [55]. Briefly, we took the Hilbert envelope of the signal filtered between 0.5 and 4.5 Hz, and divided this by its mean value to remove absolute amplitude differences. We next took the derivative of this signal – a timeseries with normally distributed values centred on zero. The TVI was defined as the standard deviation of these timeseries values.

### Quantifying the diurnal pattern of SWA

To quantify the time towards which SWA was biased at a patient level, we averaged the SWA power of both hemispheres, unless one of the SCCs was excluded from analysis (in which case the SWA estimate was based solely on the included hemisphere). SWA power estimates were log-transformed (base 10) after adding a constant of 1 to avoid zero or negative values, and then normalised to their median. This transform reduced the right-tailedness of the distribution of SWA power estimates and made it more amenable to fitting and visualisation.

To generate a fit to estimate SWA density across the 24h of the diurnal cycle, we converted the time of day associated with each of these SWA power samples to a numeric x value between 0 (00:00) and 24 (24:00). X values and associated SWA power estimates were duplicated with an offset of −24 to enable smooth fitting around midnight (00:00, x =0). This allowed us to perform a smoothing spline fit to generate an SWA estimate for each time of day. The time-of-day fit was generated using MATLAB’s *fit()* function using a smoothing spline fit type with a smoothing parameter of 0.9. To quantify the size and the timing of the most concentrated night-time SWA activity, we found the time and the value of the largest peak in the estimate between 18:00 in the evening and 10:00 in the morning (the fit segment for −6 < x < 10).

### Resampling control

LFPs were more sparsely sampled in the Pre-DBS phase (typical sampling frequency 1 LFP / 3.06 hours; Supplementary Figures S2-S10, panel A) compared to the Post-DBS phase (typical sampling frequency 1 LFP / 0.375 hours; Supplementary Figures S2-S10, panel B), which might have had an effect on the SWA power time-of-day fit and our estimates of the night-time SWA peak. We therefore investigated whether resampling the Post-DBS phase according to the Pre-DBS sample numbers and times of day would be expected to substantially alter our estimates of SWA timing. For 1000 iterations, we generated a new set of Post-DBS power estimates in the following way: For each unique time-of-day at which any Pre-DBS samples were taken, we determined the number of Pre-DBS samples taken at that exact time of day, and then randomly selected the same number of Post-DBS LFPs that were collected within half an hour on either side of this clock time. We computed an SWA time-of-day fit as described above for each of these resampled data sets, and determined its night-time peak and peak time. This resulted in a distribution of 1000 fits, 1000 peak values and 1000 peak times. For visual comparison, we compared the mean of these 1000 fits to the Pre-DBS fit (Figure S1G). For the comparison of SWA density and timing, we compared the median peak height and median peak time from the Post-DBS period with the peak height and time from the original Pre-DBS fit (Figure S1H,I). The resampled Post-DBS SWA fits for each patient and the resulting distribution of peak estimates are shown in Supplementary Figures S2-S10, panels G&H.

### Spindle detection and analysis

Spindle detection was carried out using a simplified, frequency-specific version of the spindle detection approach described by Purcell and colleagues [50]. For each hemisphere, high-SWA LFPs (>80^th^ percentile of SWA power) during the night (21:00-09:00) were selected as putative night-time NREM sleep episodes. For each high-SWA LFP, a global wavelet spectrum was calculated as follows: a continuous wavelet transform was carried out using MATLAB’s *cwt()* function with a Morse wavelet. The resulting complex transformed data was squared and averaged across the time dimension to result in a wavelet-based estimate of power at each frequency. The resulting vector was multiplied by frequency squared, which brought out a clear peak in the spindle frequency range for most data sets. The frequency corresponding to the peak in the average vector across patients and hemispheres (11.66Hz) was then used to set the centre frequency of a 7-cycle complex Morlet wavelet.

To detect spindles, each high-SWA LFP segment was convolved with this complex Morlet wavelet. Spindle power was defined as the absolute value of the complex result. We used the median absolute deviation (MAD) as a measure of variability of this signal. Any peak in the spindle power signal that exceeded median power + 5 * MAD was considered a putative spindle. For each of these putative spindles, onset was calculated as the last crossing of the lower threshold value, median spindle power + 1 * MAD, before the peak. Similarly, the spindle offset was the first time the spindle power crossed this lower threshold in the negative direction. Onset and offset values were then used to calculate spindle duration – any putative spindle with a duration under 0.5s was rejected. For each spindle, amplitude was calculated as the maximum spindle power value attained between spindle onset and offset. Spindle density was calculated as the number of spindles observed divided by the cumulative length of time of all eligible (i.e. high-SWA) LFP segments, and expressed as number of spindles per minute (assessed across all relevant 10-second LFP segments).

### Statistical analysis

Data were analysed in MATLAB R2021B (The Mathworks, Natick, MA, USA). Summary data are reported as mean ± standard deviation. For comparison of two means, both samples were tested for normality using the Shapiro-Wilk test (using A. BenSaida’s MATLAB implementation *swtest()*, revision 3.0, 2014). If no samples significantly (α < 0.05) deviated from a normal distribution, means were compared using two-tailed t-tests (paired or unpaired as appropriate). If normality was violated, medians were compared using the Wilcoxon signed-rank or Wilcoxon rank-sum tests.

## Results

### Slow-wave activity can be quantified using on-device LFP recordings from an SCC DBS implant

10-second LFP segments were obtained at regular intervals from the SCC of 9 patients implanted with bilateral DBS leads before and after a 6-month period of continuous high frequency DBS (Figure 1A-C). Slow wave activity (SWA, 0.5-4.5Hz) power was calculated for each LFP segment (Figure 1D). LFPs with SWA power above the 80^th^ percentile were classed as ‘high SWA power’ episodes that might reflect NREM sleep (Figure 1E). In these LFPs, the increase in low-frequency LFP power was specific to the lower frequencies and in particular the delta band (e.g. Figure 1F), i.e. it did not reflect a broad-band or otherwise non-specific increase in overall signal power.

While sleep has local components on shorter time scales, it is a global brain state with high levels of brain-wide synchronization, and SWA power is therefore expected to correlate strongly between 10-second LFP samples across hemispheres. To confirm this, we quantified the correlation of left and right SWA power (Figure 1G; individual participant correlations provided in Figures S2-10C,D). In the patients where both hemispheres met inclusion criteria (n = 6), correlation of left and right log SWA power (Pearson’s) was r = 0.80±0.09 (Pre-DBS) and r = 0.78±0.11 (Post-DBS; Figure 1H). The difference between the Pre-DBS and Post-DBS phase was not significant (signed-rank = 17, p = 0.219, n = 6). SWA power correlations between hemispheres for all patients are visualised as scatter plots in Supplementary Figures S2-S10, C&D.

### Slow-wave activity is more temporally focused and occurs earlier in the night after successful SCC DBS for depression

We characterised SWA power across the 24 hours of the day during the Pre- and Post-DBS periods using a smoothing spline fit to SWA power in the LFP segments corresponding to each time of day (Figure 2A-F). This revealed that, across patients, there was an earlier and sharper increase in SWA power at night in the Post-DBS period (Figure 2G). We quantified the size as well as the timing of the greatest night-time (18:00-10:00) peak of the SWA fit, and found that across patients, the SWA fit peak was significantly greater in the Post-DBS phase (Figure 2H; Pre-DBS: 2.00±0.53, Post-DBS: 2.41±0.40, t(7) = −3.81, p = 0.0066, n = 8) and occurred significantly earlier in the night (Figure 2I; 03:34±02:08, Post-DBS: 01:19±01:33, t(7) = 3.26, p = 0.0139, n = 8). The Pre-DBS and Post-DBS SWA data points and fits are provided for all patients in Supplementary Figures S2-S10, E&F. As LFPs were less frequently and less densely sampled in the Pre-DBS phase compared to the Post-DBS phase, we replicated this analysis using a random resampling approach during which Post-DBS was only sampled according to the sampling clock times and frequency from the Pre-DBS period. We generated 1000 resampled data sets per patient, and computed 1000 SWA power time-of-day fits with corresponding fit peaks (Figure S1A-G). Comparing the median of each patient’s Post-DBS fit peak distribution with the Pre-DBS fit peak still yielded a significant difference in both SWA peak size and timing (Figure S1 H,I; Peak size: Pre-DBS: 2.00±0.53, Post-DBS: 2.19±0.50, t(7) = −2.86, p = 0.024, n = 8; Peak time in 24h clock time: Pre-DBS: 03:34±02:08, Post-DBS: 01:49±01:13, t(7) = 2.69, p = 0.031, n = 8). Each patient’s distribution of Post-DBS fits and fit peak estimates is visualised in Supplementary Figures S2-S10 G&H.

**Figure 2:**
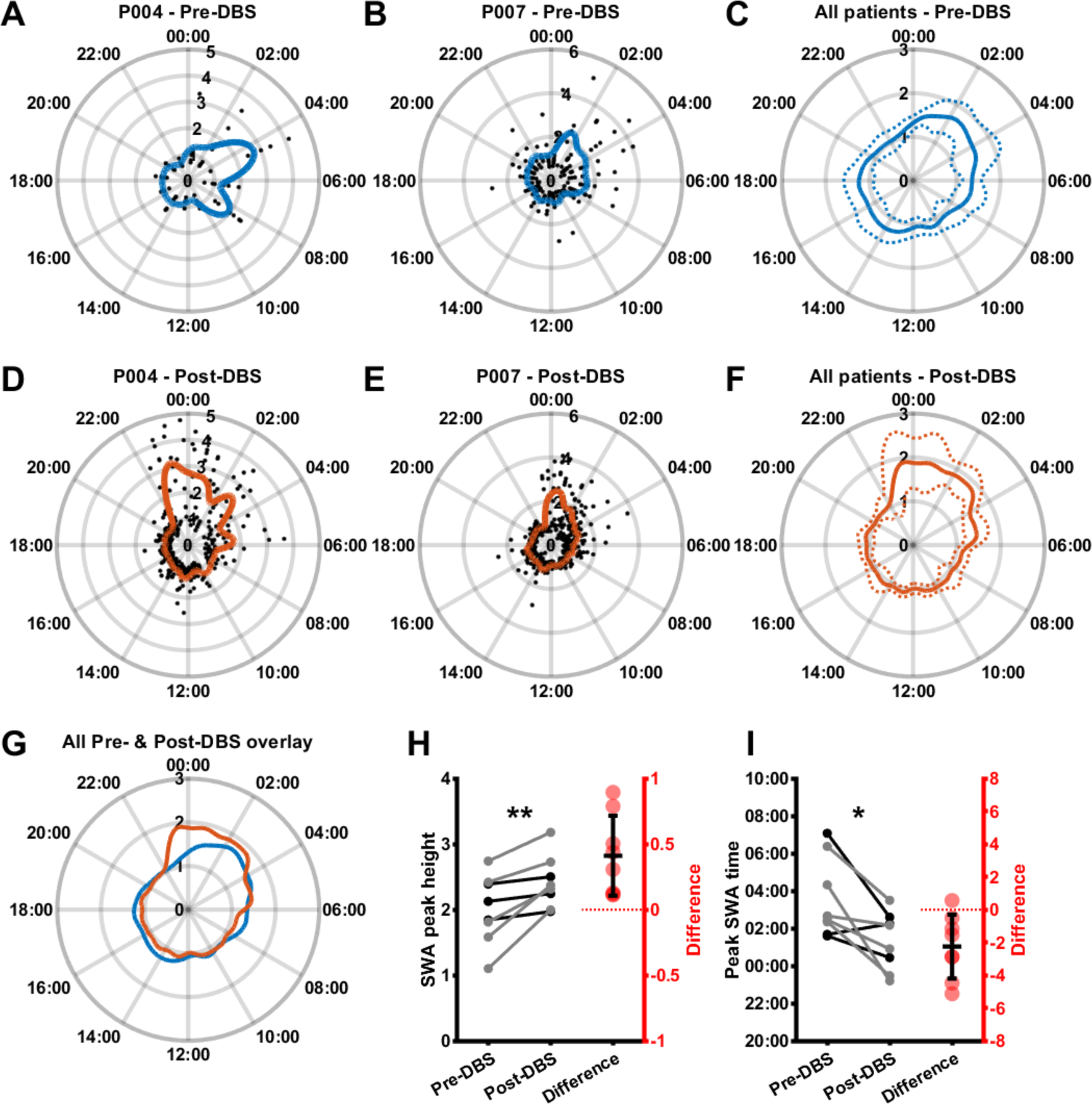
Slow-wave activity is more temporally consistent and earlier in the night after SCC DBS. **A, B:** Log-transformed and median-normalized SWA power (mean across both hemispheres; black) and a smoothing spline fit (blue) around the 24 hours of the day in the Pre-DBS phase for two example patients. **C:** Mean (solid line) plus and minus standard deviation (dotted lines) across patients of Pre-DBS time-of-day fits as illustrated in A and B. **D, E:** Log-transformed and median-normalized SWA power (mean across both hemispheres; black) and a smoothing spline fit (orange) around the 24h of the day in the Post-DBS phase for the same example patients shown in A and B. **F:** Mean (solid line) plus and minus standard deviation (dotted lines) across patients of Post-DBS time-of-day fits as illustrated in D and E. **G:** Mean Pre-DBS (blue line) and Post-DBS (orange line) time-of-day fits across patients highlighting increased SWA in the late evening and early night in the Post-DBS phase. **H:** Pre-DBS vs. Post-DBS maximum value of the SWA time-of-day fit (normalised to median fit value; Pre-DBS: 2.00±0.53, Post-DBS: 2.41±0.40, t(7) = −3.81, p = 0.0066, n = 8). Gray lines represent female patients, black lines represent male patients, red dots indicate differences. **I:** Pre-DBS vs. Post-DBS peak time (i.e. the clock time associated with the maximum value of the SWA time-of-day fit; Pre-DBS: 03:34±02:08, Post-DBS: 01:19±01:33, t(7) = 3.26, p = 0.0139, n = 8). Gray lines represent female patients, black lines represent male patients, red dots indicate differences.

**Figure 3:**
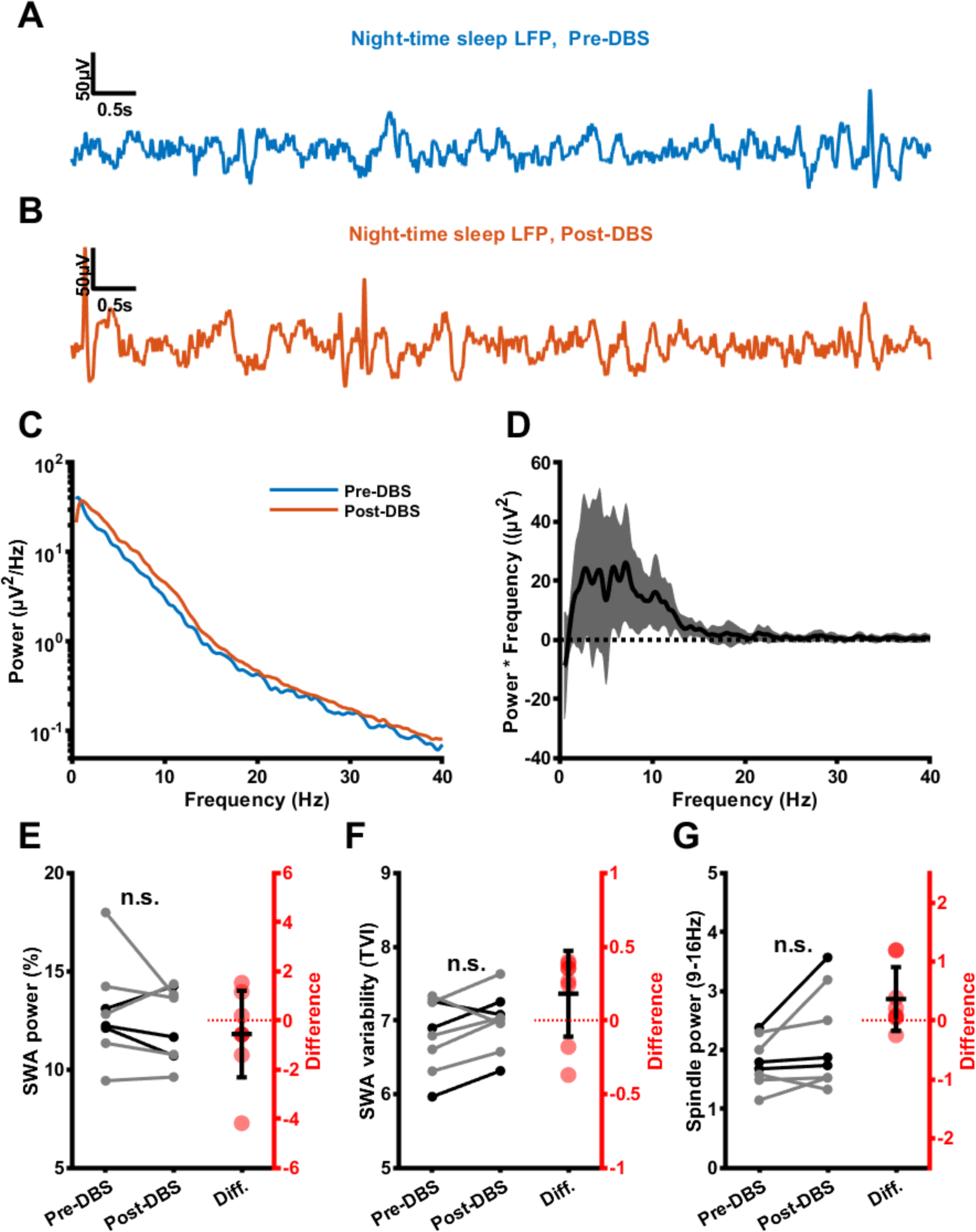
Power and stability of night-time SCC slow wave activity Pre- and Post-DBS. **A:** SCC night-time sleep LFP segment from an example patient during the Pre-DBS phase (low-pass filtered with a cutoff of 30Hz). **B:** SCC night-time sleep LFP segment from the same example patient during the Post-DBS phase (low-pass filtered with a cutoff of 30Hz). **C:** Mean night-time LFP power spectra across patients Pre-DBS (blue) and Post-DBS (orange). **D:** Mean (black line) and standard deviation (grey shaded region) night-time sleep LFP power spectral differences (multiplied by frequency for visualization) between the Pre-DBS and Post-DBS phases. **E:** Relative SWA power of night-time SCC LFPs before and after DBS (Pre-DBS: 12.9±2.49%, Post-DBS: 12.3±1.88%, t(7) = 0.895, p = 0.4, n = 8). Gray lines represent female patients, black lines represent male patients, red dots indicate differences. **F:** Temporal variability index (TVI) of night-time SCC LFPs before and after DBS (Pre-DBS: 6.8±0.489, Post-DBS: 6.98±0.4, t(7) = −1.75, p = 0.123, n = 8). Gray lines represent female patients, black lines represent male patients, red dots indicate differences. **G:** Relative spindle power of night time SCC LFPs before and after DBS (Pre-DBS: 1.79±0.415, Post-DBS: 2.15±0.837, t(7) = −1.89, p = 0.1, n = 8). Gray lines represent female patients, black lines represent male patients, red dots indicate differences.

We next compared these SWA sleep metrics to patients self-reported sleep ratings from the HDRS. Interestingly, patients already scored relatively low on the individual sleep-related items in the Pre-DBS phase, and as such there was no clear difference in scores on these items Post-DBS (see Table 2). In addition, initial SWA peak height and peak timing did not significantly correlate with Pre-DBS Hamilton score or sleep sub-scores, and changes in SWA peak height and peak timing did not significantly correlate with changes in the Hamilton overall score or sleep sub-scores (Supplementary Tables 2&3).

**Table 2:** Hamilton sleep item scores Pre-DBS vs. Post-DBS.

| HDRS item | Description | Pre-DBS | Post-DBS | p-value | signed-rank |
| --- | --- | --- | --- | --- | --- |
| HDRS4 | Difficulty falling asleep | 1.06±0.78 | 0.88±0.64 | 0.625 | 7 |
| HDRS5 | Restless and waking up in the night | 0.25±0.27 | 0.63±0.92 | 0.5 | 4 |
| HDRS6 | Early waking, unable to go back to sleep | 0.50±0.66 | 0.25±0.46 | 0.125 | 10 |
| HDRS26 | Hypersomnia | 0.31±0.37 | 0.00±0.00 | 0.125 | 10 |

### Power and stability of slow-wave activity are not altered after SCC DBS for depression

We next investigated whether the nature of SWA during putative sleep episodes was different. To exclude circadian and light cycle effects, only night-time LFPs with relatively high SWA amplitudes (LFPs < 80^th^ percentile of SWA power collected after 9pm and before 9am) were included in this analysis. Figure 3A&B show representative segments from the Pre-DBS and Post-DBS phases. We compared spectral power in these LFPs, which showed a relatively broad-band increase in the low frequencies between the Pre-DBS and Post-DBS phases (Figure 3C,D). To exclude broad-band effects resulting from any changes to the electrode-brain interface during the 6 months of DBS, we expressed power as a percentage of total signal power determined from the power spectrum. This revealed no difference in relative SWA power after SCC DBS (Figure 3e; Pre-DBS: 12.9±2.49%, Post-DBS: 12.3±1.88%, t(7) = 0.895, p = 0.4, n = 8 patients), though there was an increase in absolute power (Pre-DBS: 179±82µV^2^/Hz, Post-DBS: 227±111µV^2^/Hz, t(7) = −2.92, p = 0.0223, n = 8 patients – data not shown). We quantified the stability of slow-wave activity using a temporal variability index (TVI) that quantifies fluctuations in power envelope but is insensitive to overall changes in power [55]; see Supplementary Methods). The TVI showed no consistent difference in SWA stability between Pre-DBS and Post-DBS phases (Figure 3F; Pre-DBS: 6.8±0.489, Post-DBS: 6.98±0.4, t(7) = −1.75, p = 0.123, n = 8 patients). Finally, we compared relative power in the sleep spindle frequency range (9-16Hz), which revealed no significant increase in relative spindle power after DBS (Figure 3G; Pre-DBS: 1.79±0.415, Post-DBS: 2.15±0.837, t(7) = - 1.89, p = 0.1, n = 8 patients). However, an increase in spindle power was present in absolute terms (Pre-DBS: 25.5±16.4 µV^2^/Hz, Post-DBS: 37.4±22.5µV^2^/Hz, signed-rank = 0, p = 0.00781, n = 8 patients– data not shown).

### Sleep spindle activity is increased after successful SCC DBS treatment for depression

To investigate treatment effects on sleep spindles in more detail, we used a wavelet-based spindle detection approach (see Methods). Figure 4A shows a single night-time LFP with high SWA power from a single patient during the Post-DBS phase. Night-time SWA LFPs for this patient Post-DBS showed a peak in the frequency-adjusted wavelet spectrum in the spindle range (Figure 4B). A clear peak was present for most patients, with a mean spindle peak value of 11.66Hz, consistent with frequencies observed in cingulate cortex from previous human intracranial recordings [51]. This mean frequency was used to set the frequency of a complex 7-cycle Morlet wavelet for a wavelet transform to detect spindle events (see Methods), which successfully identified events consistent with sleep spindles (Figure 4C,D). The timing of detected spindles showed strong overlap with SWA power (e.g. Figure 4E,F), and for most patients, there was a strong correlation between times of high SWA power and times of high spindle activity (Figure 4G; Pre DBS: r = 0.414±0.385, Post DBS: r = 0.662±0.165). Moreover, we observed a shift in spindle activity between the Pre-DBS and Post-DBS phase (Figure 4H), analogous to the shift in SWA activity illustrated in Figure 2. Figure 4I shows the correlation between SWA power and percentage of spindle activity across all LFP snippets from all hemispheres (r = 0.43, p = 4.1*10^-241^). There was an increased density of spindles in the Post-DBS phase (Figure 4J; Pre DBS: 5.51±3.82 spindles/min, Post DBS: 7.26±4 spindles/min, t(7) = - 3.77, p = 0.00702, n = 8). A comparison of the properties of individual spindles revealed no change in spindle duration (Figure 4K; Pre DBS: 0.773±0.165s, Post DBS: 0.78±0.0639s, t(7) = −0.13, p = 0.901, n = 8) or amplitude (Figure 4L, Pre DBS: 4.25±0.587, Post DBS: 4.51±0.542, t(7) = −2.17, p = 0.0668, n = 8). Changes in spindle density did not significantly correlate with changes in the Hamilton overall score or sleep items (Supplementary Table 4).

**Figure 4:**
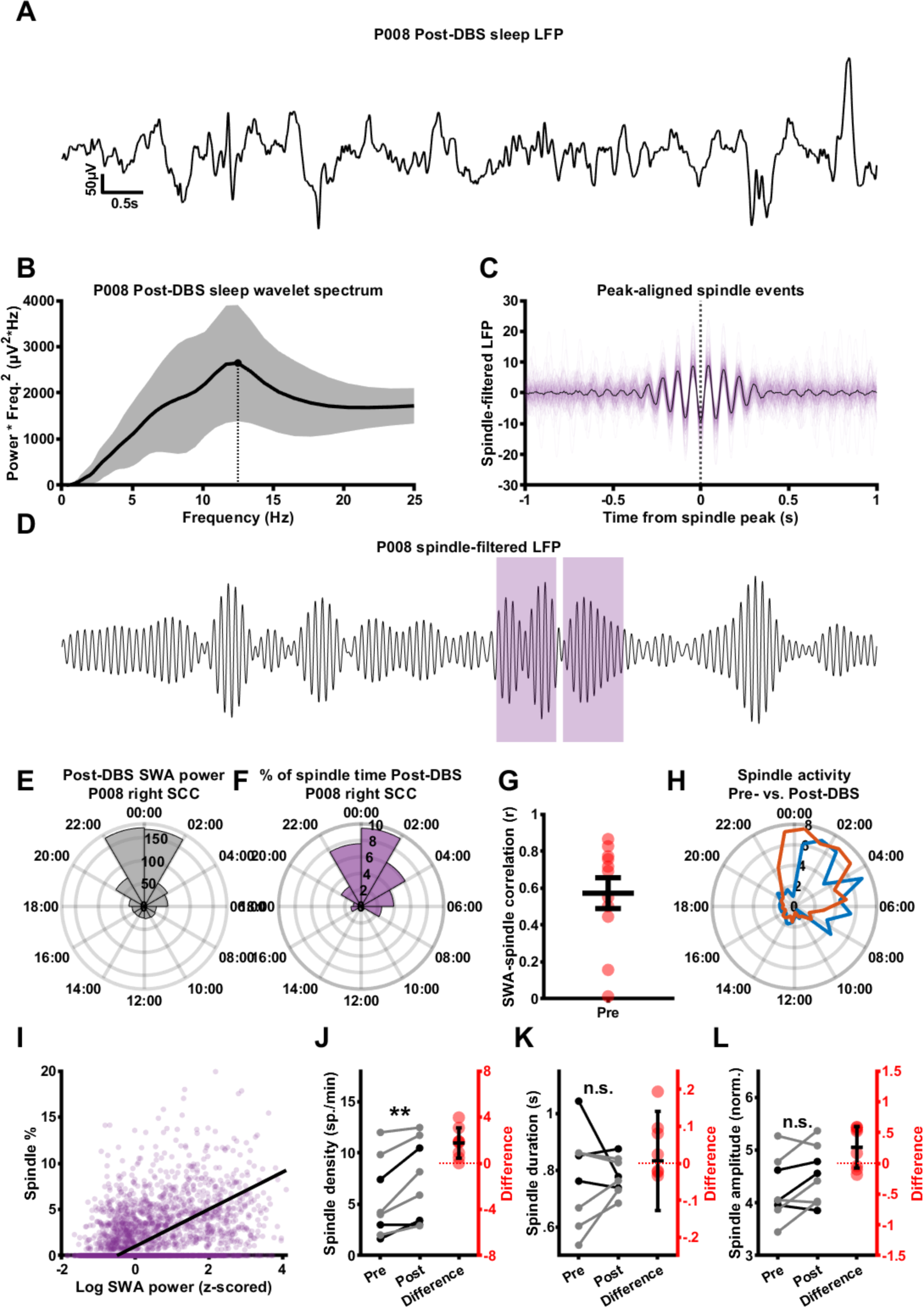
Sleep spindle activity is increased in the post-DBS phase. **A:** A night-time sleep LFP segment from the right SCC of an example patient, collected during the Post-DBS period (low-pass filtered with a cutoff frequency of 30Hz). **B:** The mean (black line) and standard deviation (grey area) of the global wavelet spectra of all Post-DBS night-time sleep LFP segments for this example hemisphere, multiplied by frequency^2^ to bring out the spindle peak. The point and dotted line indicate the spindle peak for this particular patient. **C:** All spindle events detected in night-time sleep LFP segments from the Post-DBS phase for this hemisphere, aligned to the spindle peak (transparent orange lines) and their mean (black line). **D:** The same LFP segment as in A, narrow-band filtered around the spindle peak frequency (11-12 Hz pass band), with orange sections indicating detected spindle events. **E:** Median night-time SWA power for this example hemisphere at different times of day for this example hemisphere. **F:** Percentage of spindle activity across LFP segments collected at different times of day for this example hemisphere. Note the close association with SWA power in E. **G:** Pearson’s correlation coefficients of SWA power and spindle percentage for the Pre-DBS and Post-DBS periods Pre DBS: r = 0.414±0.385, Post DBS: r = 0.662±0.165. **H:** Mean percentage of spindle activity across patients in the Pre-DBS (blue) and Post-DBS (orange) period across the 24h diurnal cycle. Note the shift towards earlier spindle times in the Post-DBS period. **I:** Scatter plot of log10 slow wave power vs. spindle percentage for all individual LFP snippets in the Post-DBS data set across participants, with regression line (purple) showing a clear positive correlation. **J:** Paired line plot representing spindle density before and after SCC DBS (Pre DBS: 5.51±3.82 spindles/min, Post DBS: 7.26±4 spindles/min, t(7) = −3.77, p = 0.00702, n = 8). Gray lines represent female patients, black lines represent male patients, red dots indicate differences. **K:** Paired line plot representing spindle duration before and after SCC DBS (Pre DBS: 0.773±0.165s, Post DBS: 0.78±0.0639s, t(7) = −0.13, p = 0.901, n = 8). Gray lines represent female patients, black lines represent male patients, red dots indicate differences. **L:** Paired line plot representing spindle amplitude before and after SCC DBS (Pre DBS: 4.25±0.587, Post DBS: 4.51±0.542, t(7) = −2.17, p = 0.0668, n = 8). Gray lines represent female patients, black lines represent male patients, red dots indicate differences.

## Discussion

This study characterised changes in intracortical sleep signatures following 24 weeks of DBS treatment for depression. We demonstrate that the timing of SWA shifts towards earlier hours of the night after clinically effective high-frequency SCC DBS. While the overall power and stability of SWA did not appear different between the Pre-DBS and Post-DBS study phases, the density of sleep spindle activity increased. To the best of our knowledge, this is the first study to report electrophysiological signatures of sleep in SCC and changes in these signatures and sleep-wake cycle following SCC DBS. While there are several caveats to interpreting these findings in the context of treatment, the combination of recording and analysis used here provides a novel approach to characterising the modulation of sleep-related brain activity by DBS in depression and other brain disorders.

The timeline of the trial dictated that recordings that stimulation-off/artifact-free recordings were made for 1 month after surgery (Pre-DBS) and for 1 week following a period of 6 months of 130Hz stimulation (Post-DBS). Bearing in mind that the improvement in depressive symptoms remained stable for all patients during the blinded discontinuation Post-DBS week, there are important caveats for interpreting any changes between these phases. Firstly, this timeline does not allow us to distinguish between 1) changes due directly to DBS that persist and are unrelated to depressive symptoms, 2) changes due to the effect of DBS on depressive symptoms or 3) changes due to effect of turning DBS off after a prolonged period of stimulation. Secondly, the patients’ routine and the data collection frequency in the Pre-DBS and Post-DBS phases was different, as the patients attended the clinic every day in the Post-DBS phase. Finally, as all the patients in this cohort had significant improvement in depressive symptoms, we could not use variance in clinical response to delineate direct effects on sleep and secondary effects of symptom improvement. . We will consider these caveats carefully when interpreting our results.

Our first important observation is a significant shift in SWA towards the earlier part of the night after DBS. Note that using the peak of the time-of-day fit to estimate peak SWA density does not distinguish between 1) higher SWA power or 2) a greater proportion of high-SWA (e.g. NREM sleep) during a particular period. Either of these scenarios is consistent with multiple explanations, for instance: a reduced difficulty falling asleep at night, a change in sleep structure favouring deeper NREM stages in the early sleep cycles, or a shift in circadian pattern towards an earlier chronotype (these explanations are not mutually exclusive). Secondly, using a wavelet-based spectrum, we found a peak sleep spindle frequency of 11-12 Hz. This is consistent with spindles observed at human frontal EEG sites [50,56], and matches intracranial data from the adjacent human anterior cingulate cortex [51]. We show that spindle density is increased in the Post-DBS period. In contrast to the change in SWA profile, study-related changes in the patient’s routine are relatively unlikely to result in changes in spindle density. As spindles occur most frequently in NREM stage 2, one possibility is that patients spend more time in NREM stage 2 sleep in the Post-DBS phase. If this were the case, a change in slow wave power and/or stability would also be expected, but we found no evidence for this.

Overall, our findings are consistent with the observation that earlier, more consistent sleep patterns across nights [7–18,33] and a sleep architecture that favours the earlier occurrence of deeper NREM sleep stages [19–27] are associated with good outcomes in the treatment of depression. In previous reports on SCC DBS, sleep was one of the first symptoms to show improvement with treatment. Mayberg et al. reported that DBS improved early-morning sleep disturbance in responders while not affecting difficulty falling asleep and hypersomnia seen in the nonresponders [1]. Lozano et al. further reported an improvement in the sleep subscore of HDRS after 1 month of DBS that persisted over 12 months [2]. In this cohort, we did not find improvements on HDRS sleep scores between Pre- and Post-DBS, and no correlation between the changes in electrophysiological markers and specific sleep items of the Hamilton depression rating scale, or the overall Hamilton score. A potential reason is that the characterisation of sleep on the Hamiliton scale has a limited range and variability, with the individual items allowing for 3 possible ratings (0,1,2). Overall, in a cohort of 9 patients with the same direction of response, we had limited statistical power to establish an association between our sleep metrics and subjective reports. Our results highlight that studies in a larger cohort are warranted, and these should consider more elaborate sleep self-reporting as well as the use of wearable activity trackers for more objective sleep time estimates. Importantly, several of our patients were taking benzodiazepines either as needed or at bedtime, both pre- and post-DBS. This could partly account for the low incidence of subjective sleep disturbances in these patients [57].

What mechanisms could underlie changes in SWA and sleep spindles following SCC DBS? SWA and spindle activities result from the complex interplay of cortico-thalamic circuits under the regulation of ascending afferents from brainstem nuclei [58]. DBS has been demonstrated to affect SCC LFPs through modulation of the local circuit [40,52,54,59], but because of the global nature of these sleep-related oscillations, it is more likely that the observed changes are due to effects on the network. The anatomy of the SCC supports this conjecture, as the SCC acts as a hub in the limbic circuit, projecting to and receiving projections from many other regions implicated in sleep [60]. For example, animal studies have shown that SCC projects to dorsal raphe nuclei (DRN) [61], a major serotonergic region of the brain, and alters the firing patterns of serotonergic neurons [62,63]. Serotonin has been linked to the sleep-wake cycle, [64,65], and serotonergic medications such as SSRIs have a known effect on sleep structure, in particular suppressing and delaying episodes of REM sleep [66]. SCC also projects to the hypothalamus [67], a key region controlling arousal and sleep-wake cycles that contains the suprachiasmatic nucleus, our ‘circadian master clock’ [68,69]. SCC has also been shown to moderately to weakly project to the thalamic reticular nuclei [67], which are currently considered the originators of sleep spindles [48,49]. SCC DBS therefore has several plausible ways of influencing cortical signatures of sleep that merit further investigation.

### Quality control and limitations

Recording LFPs regularly over long periods provides a unique opportunity characterise brain activity associated with healthy and disease-related activity and their modulation by therapies. However, as we and others have previously shown, obtaining such recordings while patients go about their daily lives provides several sources of possible artifact [43,70]. While the majority of the data sets were of sufficient quality for analysing sleep-related activity, data from a number of hemispheres had to be excluded on the basis of ECG artifact or amplifier saturation. One simple quality control measure we used was to look at the correlation of SWA across hemispheres, as during NREM sleep, SWA is found across the entire forebrain [58]. We found SWA in SCC LFPs to be consistent with global synchronisation, as the power of SWA was highly correlated across hemispheres. Both the temporal profile and synchronisation of SWA power were consistent with accurate detection of these activities, though we acknowledge that ground truth polysomnographic sleep scoring would be necessary to conclusively verify this. The sampling density of LFP data in the Pre- and Post-DBS phases was determined based on the storage capacity of the device and the time period between in-clinic visits that would allow for the downloading of the data. Moreover, the timing of some clinical visits themselves led to some gaps in the sampling of the data (as shown in Supplementary Figures S2-S10 for individual patients). As this might have particularly influenced our estimates of SWA timing, we controlled for this by using a resampling approach, which tested the consistency of the Post-DBS SWA fits and metrics with a sparser sampling regime. While it would be ideal for sampling to be consistent between patients and phases, datasets recorded over weeks away from the clinic will often require sparse sampling to preserve battery life. Importantly, we demonstrate that sparse and/or irregular sampling does not prevent meaningful analysis and such datasets remain highly valuable. Moreover, this approach allows data to be recorded with minimal disruption to the patient’s routine by visits to the clinic. Next-generation DBS sensing devices[71–73] may allow more controlled night recordings to replicate and extend these initial observations.

In the Pre-DBS phase, we only included data acquired at least 14 days post-surgery, to minimize the impact of surgical recovery on our analysis. However, it is likely that after-effects of neurosurgery (whether behavioural or neurophysiological) still had an impact on our Pre-DBS results. Specifically, during this time, there was already a significant improvement in depressive symptoms, which could indicate the presence of a lesion effect at this time [53]. Secondly, the requirement to come into the clinic daily during the Post-DBS phase, as noted, could have had an effect on patient behavioural routine and therefore have influenced bed times and their consistency across days. It would be less likely to affect sleep architecture or sleep spindle density. Finally, it should be noted that many antidepressants [66] and benzodiazepines [57] have known effects on sleep architecture. However, despite the variance of medications used across the subjects, all subjects remained on stable doses of their pre-surgical medications throughout the study, so such medication-specific effects are not expected to underlie the consistent findings across subjects.

## Conclusion

We provide evidence that it is possible to detect cortical sleep signatures chronically recorded on-device from DBS leads in the SCC, and to investigate their characteristics and diurnal pattern. Despite limitations in the data set, it was possible to detect differences in sleep pattern and spindle activity between a pre- and post-treatment period. Further work is needed to determine whether such changes are related to SCC DBS, symptom remission, or changes in behaviour. The ability to chronically estimate sleep state from data recorded on a DBS device opens up the possibility of DBS therapy strategies that are sensitive to a patient’s diurnal behaviour and sleep/wake patterns [36,37].

## Data availability

The anonymised raw data (LFP timeseries and associated timestamps, as well as associated clinical data) are publicly available as part of the data set released for Alagapan et al. 2023 [42] via the Data Archive for the Brain Initiative (DABI) at https://dabi.loni.usc.edu/dsi/1UH3NS103550/UXUF7822Z3JL. Pre-processed data as well as MATLAB code to reproduce figures and analyses in this paper will be made available on request from the corresponding authors.

Andrew Sharott

Joram van Rheede

## Acknowledgements

This study was supported by the following grants: Hope for Depression Research Foundation, NIH UH3NS103550; Medical Research Council UK, MC_UU_00003/6 to AS and MC_UU_00003/3 to TD. The Activa PC + S DBS devices used in this study were donated by Medtronic.

## References

1. Mayberg HS, Lozano AM, Voon V, et al. Deep Brain Stimulation for Treatment-Resistant Depression. Neuron. 2005;45(5):651–660. doi:10.1016/j.neuron.2005.02.014

2. Lozano AM, Mayberg HS, Giacobbe P, Hamani C, Craddock RC, Kennedy SH. Subcallosal Cingulate Gyrus Deep Brain Stimulation for Treatment-Resistant Depression. Biol Psychiat. 2008;64(6):461–467. doi:10.1016/j.biopsych.2008.05.034

3. Holtzheimer PE, Kelley ME, Gross RE, et al. Subcallosal Cingulate Deep Brain Stimulation for Treatment-Resistant Unipolar and Bipolar Depression. Arch Gen Psychiat. 2012;69(2):150–158. doi:10.1001/archgenpsychiatry.2011.1456

4. Riva-Posse P, Choi KS, Holtzheimer PE, et al. A connectomic approach for subcallosal cingulate deep brain stimulation surgery: prospective targeting in treatment-resistant depression. Mol Psychiatr. 2018;23(4):843–849. doi:10.1038/mp.2017.59

5. Brown EC, Clark DL, Forkert ND, Molnar CP, Kiss ZHT, Ramasubbu R. Metabolic activity in subcallosal cingulate predicts response to deep brain stimulation for depression. Neuropsychopharmacol. 2020;45(10):1681–1688. doi:10.1038/s41386-020-0745-5

6. American Psychiatric Association. Diagnostic and Statistical Manual of Mental Disorders (5th Ed.) - Text Revision.; 2022.

7. Mirchandaney R, Asarnow LD, Kaplan KA. Recent advances in sleep and depression. Curr Opin Psychiatr. 2023;36(1):34–40. doi:10.1097/yco.0000000000000837

8. Tsuno N, Besset A, Ritchie K. Sleep and Depression. J Clin Psychiatry. 2005;66(10):1254–1269. doi:10.4088/jcp.v66n1008

9. Riemann D, Berger M, Voderholzer U. Sleep and depression — results from psychobiological studies: an overview. Biol Psychol. 2001;57(1-3):67–103. doi:10.1016/s0301-0511(01)00090-4

10. Alvaro PK, Roberts RM, Harris JK. A Systematic Review Assessing Bidirectionality between Sleep Disturbances, Anxiety, and Depression. Sleep. 2013;36(7):1059–1068. doi:10.5665/sleep.2810

11. Fang H, Tu S, Sheng J, Shao A. Depression in sleep disturbance: A review on a bidirectional relationship, mechanisms and treatment. J Cell Mol Med. 2019;23(4):2324–2332. doi:10.1111/jcmm.14170

12. Taylor DJ, Lichstein KL, Durrence HH, Reidel BW, Bush AJ. Epidemiology of Insomnia, Depression, and Anxiety. Sleep. 2005;28(11):1457-1464. doi:10.1093/sleep/28.11.1457

13. Buysse DJ, Angst J, Gamma A, Ajdacic V, Eich D, Rössler W. Prevalence, Course, and Comorbidity of Insomnia and Depression in Young Adults. Sleep. 2008;31(4):473–480. doi:10.1093/sleep/31.4.473

14. Plante DT. The Evolving Nexus of Sleep and Depression. Am J Psychiat. 2021;178(10):896–902. doi:10.1176/appi.ajp.2021.21080821

15. Chan JWY, Lam SP, Li SX, et al. Eveningness and Insomnia: Independent Risk Factors of Nonremission in Major Depressive Disorder. Sleep. 2014;37(5):911–917. doi:10.5665/sleep.3658

16. Dombrovski AY, Mulsant BH, Houck PR, et al. Residual symptoms and recurrence during maintenance treatment of late-life depression. J Affect Disorders. 2007;103(1-3):77–82. doi:10.1016/j.jad.2007.01.020

17. Combs K, Smith PJ, Sherwood A, et al. Impact of Sleep Complaints and Depression Outcomes Among Participants in the Standard Medical Intervention and Long-Term Exercise Study of Exercise and Pharmacotherapy for Depression. J Nerv Ment Dis. 2014;202(2):167–171. doi:10.1097/nmd.0000000000000085

18. Manglick M, Rajaratnam SM, Taffe J, Tonge B, Melvin G. Persistent sleep disturbance is associated with treatment response in adolescents with depression. Australian New Zealand J Psychiatry. 2013;47(6):556–563. doi:10.1177/0004867413481630

19. Kupfer DJ, Foster FG, Reich L, Thompson SK, Weiss B. EEG sleep changes as predictors in depression. Am J Psychiatry. 1976;133(6):622–626. doi:10.1176/ajp.133.6.622

20. Kupfer David J, Foster FG. INTERVAL BETWEEN ONSET OF SLEEP AND RAPID-EYE-MOVEMENT SLEEP AS AN INDICATOR OF DEPRESSION. Lancet. 1972;300(7779):684–686. doi:10.1016/s0140-6736(72)92090-9

21. Ehlers CL, Havstad JW, Kupfer DJ. Estimation of the time course of slow-wave sleep over the night in depressed patients: effects of clomipramine and clinical response. Biol Psychiatry. 1996;39(3):171–181. doi:10.1016/0006-3223(95)00139-5

22. Kupfer DJ. Sleep research in depressive illness: Clinical implications —a tasting menu. Biol Psychiatry. 1995;38(6):391–403. doi:10.1016/0006-3223(94)00295-e

23. Benca RM, Obermeyer WH, Thisted RA, Gillin JC. Sleep and Psychiatric Disorders: A Meta-analysis. Arch Gen Psychiatry. 1992;49(8):651–668. doi:10.1001/arch-psyc.1992.01820080059010

24. Perlis ML, Giles DE, Buysse DJ, Thase ME, Tu X, Kupfer DJ. Which depressive symptoms are related to which sleep electroencephalographic variables? Biol Psychiatry. 1997;42(10):904–913. doi:10.1016/s0006-3223(96)00439-8

25. Steiger A, Kimura M. Wake and sleep EEG provide biomarkers in depression. J Psychiatr Res. 2010;44(4):242–252. doi:10.1016/j.jpsychires.2009.08.013

26. Wichniak A, Wierzbicka A, Jernajczyk W. Sleep as a biomarker for depression. Int Rev Psychiatry. 2013;25(5):632–645. doi:10.3109/09540261.2013.812067

27. Steiger A, Pawlowski M. Depression and Sleep. Int J Mol Sci. 2019;20(3):607. doi:10.3390/ijms20030607

28. Armitage R, Hoffmann R, Trivedi M, Rush AJ. Slow-wave activity in NREM sleep: sex and age effects in depressed outpatients and healthy controls. Psychiatry Res. 2000;95(3):201–213. doi:10.1016/s0165-1781(00)00178-5

29. Armitage R. Sleep and circadian rhythms in mood disorders. Acta Psychiat Scand. 2007;115(s433):104–115. doi:10.1111/j.1600-0447.2007.00968.x

30. Gaspar-Barba E, Calati R, Cruz-Fuentes CS, et al. Depressive symptomatology is influenced by chronotypes. J Affect Disorders. 2009;119(1-3):100–106. doi:10.1016/j.jad.2009.02.021

31. Antypa N, Vogelzangs N, Meesters Y, Schoevers R, Penninx BWJH. Chronotype associations with depression and anxiety disorders in a large cohort study. Dépress Anxiety. 2016;33(1):75–83. doi:10.1002/da.22422

32. Au J, Reece J. The relationship between chronotype and depressive symptoms: A meta-analysis. J Affect Disord. 2017;218:93–104. doi:10.1016/j.jad.2017.04.021

33. O’Loughlin J, Casanova F, Jones SE, et al. Using Mendelian Randomisation methods to understand whether diurnal preference is causally related to mental health. Mol Psychiatr. 2021;26(11):6305–6316. doi:10.1038/s41380-021-01157-3

34. Jagannath A, Peirson SN, Foster RG. Sleep and circadian rhythm disruption in neuro-psychiatric illness. Curr Opin Neurobiol. 2013;23(5):888–894. doi:10.1016/j.conb.2013.03.008

35. Scangos KW, Khambhati AN, Daly PM, et al. Closed-loop neuromodulation in an individual with treatment-resistant depression. Nat Med. 2021;27(10):1696–1700. doi:10.1038/s41591-021-01480-w

36. Gilron R, Little S, Wilt R, Perrone R, Anso J, Starr PA. Sleep-aware adaptive deep brain stimulation control: Chronic use at home with dual independent linear discriminate detectors. Front Neurosci-switz. 2021;15:732499. doi:10.3389/fnins.2021.732499

37. Fleming JE, Kremen V, Gilron R, et al. Embedding digital chronotherapy into bioelectronic medicines. Iscience. 2022;25(4):104028. doi:10.1016/j.isci.2022.104028

38. Stanslaski S, Herron J, Chouinard T, et al. A Chronically Implantable Neural Coprocessor for Investigating the Treatment of Neurological Disorders. Ieee T Biomed Circ S. 2018;12(6):1230–1245. doi:10.1109/tbcas.2018.2880148

39. Stanslaski S, Afshar P, Cong P, et al. Design and Validation of a Fully Implantable, Chronic, Closed-Loop Neuromodulation Device with Concurrent Sensing and Stimulation. Ieee T Neur Sys Reh. 2012;20(4):410–421. doi:10.1109/tnsre.2012.2183617

40. Veerakumar A, Tiruvadi V, Howell B, et al. Field potential 1/f activity in the subcallosal cingulate region as a candidate signal for monitoring deep brain stimulation for treatment-resistant depression. J Neurophysiol. 2019;122(3):1023–1035. doi:10.1152/jn.00875.2018

41. Tiruvadi VR, Choi KS, Waters A, et al. Network Action of Subcallosal Cingulate White Matter Deep Brain Stimulation. Medrxiv. Published online 2022:2022.07.27.22278130. doi:10.1101/2022.07.27.22278130

42. Alagapan S, Choi KS, Heisig S, et al. Cingulate dynamics track depression recovery with deep brain stimulation. Nature. 2023;622(7981):130-138. doi:10.1038/s41586-023-06541-3

43. Rheede JJ van, Feldmann LK, Busch JL, et al. Diurnal modulation of subthalamic beta oscillatory power in Parkinson’s disease patients during deep brain stimulation. Npj Park Dis. 2022;8(1):88. doi:10.1038/s41531-022-00350-7

44. Steriade M, McCormick DA, Sejnowski TJ. Thalamocortical oscillations in the sleeping and aroused brain. Science. 1993;262(5134):679–685. doi:10.1126/science.8235588

45. Carskadon MA, Dement WC. Principles and Practice of Sleep Medicine (Fifth Edition). Part Princ Sleep Medicine Sect 1normal Sleep Var Sect 1 Normal Sleep Var. Published online 2011:16–26. doi:10.1016/b978-1-4160-6645-3.00002-5

46. Lüthi A. Sleep Spindles. Neurosci. 2014;20(3):243–256. doi:10.1177/1073858413500854

47. Jankel WR, Niedermeyer E. Sleep Spindles. J Clin Neurophysiol. 1985;2(1):1-36. doi:10.1097/00004691-198501000-00001

48. Fernandez LMJ, Lüthi A. Sleep Spindles: Mechanisms and Functions. Physiol Rev. 2020;100(2):805–868. doi:10.1152/physrev.00042.2018

49. Gennaro LD, Ferrara M. Sleep spindles: an overview. Sleep Med Rev. 2003;7(5):423–440. doi:10.1053/smrv.2002.0252

50. Purcell SM, Manoach DS, Demanuele C, et al. Characterizing sleep spindles in 11,630 individuals from the National Sleep Research Resource. Nat Commun. 2017;8(1):15930. doi:10.1038/ncomms15930

51. Andrillon T, Nir Y, Staba RJ, et al. Sleep Spindles in Humans: Insights from Intracranial EEG and Unit Recordings. J Neurosci. 2011;31(49):17821–17834. doi:10.1523/jneurosci.2604-11.2011

52. Sendi MSE, Waters AC, Tiruvadi V, et al. Intraoperative neural signals predict rapid anti-depressant effects of deep brain stimulation. Transl Psychiat. 2021;11(1):551. doi:10.1038/s41398-021-01669-0

53. Mestre TA, Lang AE, Okun MS. Factors influencing the outcome of deep brain stimulation: Placebo, nocebo, lessebo, and lesion effects. Mov Disord. 2016;31(3):290–298. doi:10.1002/mds.26500

54. Alagapan S, Choi K, Heisig S, et al. Cingulate dynamics track depression recovery with deep brain stimulation. Nature.

55. McNamara CG, Rothwell M, Sharott A. Stable, interactive modulation of neuronal oscillations produced through brain-machine equilibrium. Cell Reports. 2022;41(6):111616. doi:10.1016/j.celrep.2022.111616

56. Plante DT, Goldstein MR, Landsness EC, et al. Topographic and sex-related differences in sleep spindles in major depressive disorder: A high-density EEG investigation. J Affect Disorders. 2013;146(1):120–125. doi:10.1016/j.jad.2012.06.016

57. Mendonça FMR de, Mendonça GPRR de, Souza LC, et al. Benzodiazepines and Sleep Architecture: A Systematic Review. CNS Neurol Disord - Drug Targets. 2023;22(2):172–179. doi:10.2174/1871527320666210618103344

58. Steriade M. Grouping of brain rhythms in corticothalamic systems. Neuroscience. 2006;137(4):1087–1106. doi:10.1016/j.neuroscience.2005.10.029

59. Smart O, Choi KS, Riva-Posse P, et al. Initial Unilateral Exposure to Deep Brain Stimulation in Treatment-Resistant Depression Patients Alters Spectral Power in the Subcallosal Cingulate. Front Comput Neurosc. 2018;12:43. doi:10.3389/fncom.2018.00043

60. Hamani C, Mayberg H, Stone S, Laxton A, Haber S, Lozano AM. The Subcallosal Cingulate Gyrus in the Context of Major Depression. Biol Psychiat. 2011;69(4):301–308. doi:10.1016/j.biopsych.2010.09.034

61. Freedman LJ, Insel TR, Smith Y. Subcortical projections of area 25 (subgenual cortex) of the macaque monkey. J Comp Neurol. 2000;421(2):172–188. doi:10.1002/(sici)1096-9861(20000529)421:2<172::aid-cne4>3.0.co;2-8

62. Srejic LR, Hamani C, Hutchison WD. High-frequency stimulation of the medial prefrontal cortex decreases cellular firing in the dorsal raphe. Eur J Neurosci. 2015;41(9):1219–1226. doi:10.1111/ejn.12856

63. Srejic LR, Wood KM, Zeqja A, Hashemi P, Hutchison WD. Modulation of serotonin dynamics in the dorsal raphe nucleus via high frequency medial prefrontal cortex stimulation. Neurobiol Dis. 2016;94:129–138. doi:10.1016/j.nbd.2016.06.009

64. Portas CM, Bjorvatn B, Ursin R. Serotonin and the sleep/wake cycle: special emphasis on microdialysis studies. Prog Neurobiol. 2000;60(1):13–35. doi:10.1016/s0301-0082(98)00097-5

65. Monti JM. Serotonin control of sleep-wake behavior. Sleep Med Rev. 2011;15(4):269–281. doi:10.1016/j.smrv.2010.11.003

66. Wichniak A, Wierzbicka A, Walęcka M, Jernajczyk W. Effects of Antidepressants on Sleep. Curr Psychiat Rep. 2017;19(9):63. doi:10.1007/s11920-017-0816-4

67. Chiba T, Kayahara T, Nakano K. Efferent projections of infralimbic and prelimbic areas of the medial prefrontal cortex in the Japanese monkey, Macaca fuscata. Brain Res. 2001;888(1):83–101. doi:10.1016/s0006-8993(00)03013-4

68. Saper CB, Scammell TE, Lu J. Hypothalamic regulation of sleep and circadian rhythms. Nature. 2005;437(7063):1257–1263. doi:10.1038/nature04284

69. Ralph MR, Foster RG, Davis FC, Menaker M. Transplanted Suprachiasmatic Nucleus Determines Circadian Period. Science. 1990;247(4945):975–978. doi:10.1126/science.2305266

70. Neumann WJ, Sorkhabi MM, Benjaber M, et al. The sensitivity of ECG contamination to surgical implantation site in brain computer interfaces. Brain Stimul. 2021;14(5):1301–1306. doi:10.1016/j.brs.2021.08.016

71. Zamora M, Toth R, Morgante F, et al. DyNeuMo Mk-1: Design and pilot validation of an investigational motion-adaptive neurostimulator with integrated chronotherapy. Exp Neurol. 2022;351:113977. doi:10.1016/j.expneurol.2022.113977

72. Toth R, Zamora M, Ottaway J, et al. DyNeuMo Mk-2: An Investigational Circadian-Locked Neuromodulator with Responsive Stimulation for Applied Chronobiology. 2020 Ieee Int Conf Syst Man Cybern Smc. 2020;00:3433–3440. doi:10.1109/smc42975.2020.9283187

73. Herron J, Stanslaski S, Chouinard T, Corey R, Denison T, Orser H. Bi-directional brain interfacing instrumentation. 2018 IEEE Int Instrum Meas Technol Conf (I2MTC). Published online 2018:1-6. doi:10.1109/i2mtc.2018.8409795

