## Supplementary Figures and Tables for "Cortical signatures of sleep are altered following effective deep brain stimulation for depression"

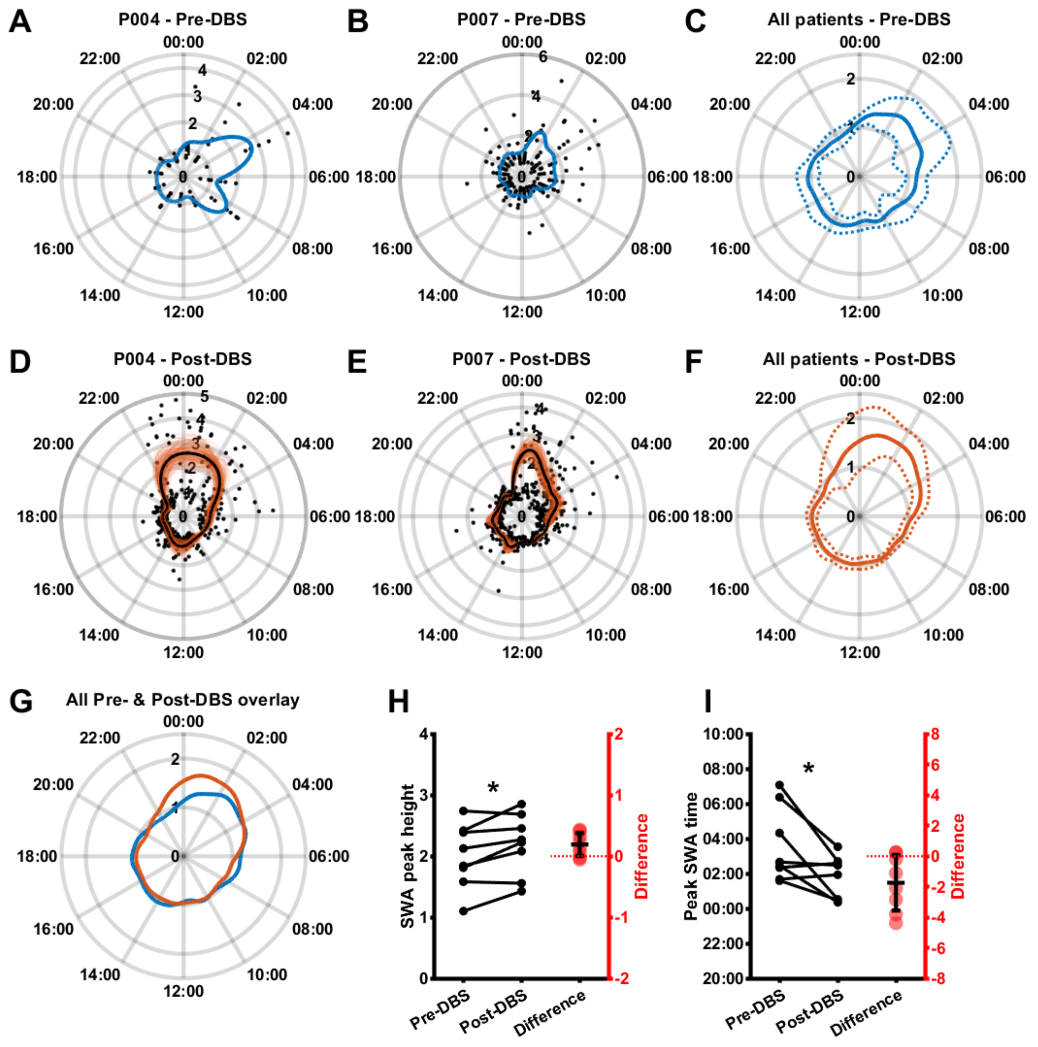

**Figure S1: Random resampling of the Post-DBS phase confirms that slow-wave activity is more temporally consistent and occurs earlier in the night after SCC DBS**

**A, B:** Log-transformed and median-normalized SWA power (mean across both hemispheres; black) and a smoothing spline fit (blue) around the 24 hours of the day in the Pre-DBS phase for two example patients. **C:** Mean (solid line) plus and minus standard deviation (dotted lines) across patients of Pre-DBS time-of-day fits as illustrated in A and B. **D, E:** Log-transformed and median-normalized SWA power (mean across both hemispheres; black) and 1000 overlaid smoothing spline fits (orange) as well as their mean (black) around the 24h of the day in the Post-DBS phase for the same example patients shown in A and B. **F:** Mean (solid line) plus and minus standard deviation (dotted lines) across patients of the mean resampled Post-DBS time-of-day fits as illustrated in D and E. **G:** Mean Pre-DBS (blue line) and resampled Post-DBS (orange line) time-of-day fits across patients highlighting increased SWA in the late evening and early night in the Post-DBS phase. **H:** Pre-DBS vs. resampled Post-DBS median height of the greatest night-time SWA peak as estimated by the time-of-day fit (normalised to median; Pre-DBS: 2.00±0.53, Post-DBS: 2.41±0.40, $t(7) = -3.81$, $p = 0.0066$, $n = 8$). **I:** Pre-DBS vs. resampled Post-DBS median time of day of the greatest night-time SWA peak as estimated by the time-of-day fit (in 24h clock time; Pre-DBS: 03:34±02:08, Post-DBS: 01:19±01:33, $t(7) = 3.26$, $p = 0.0139$, $n = 8$).

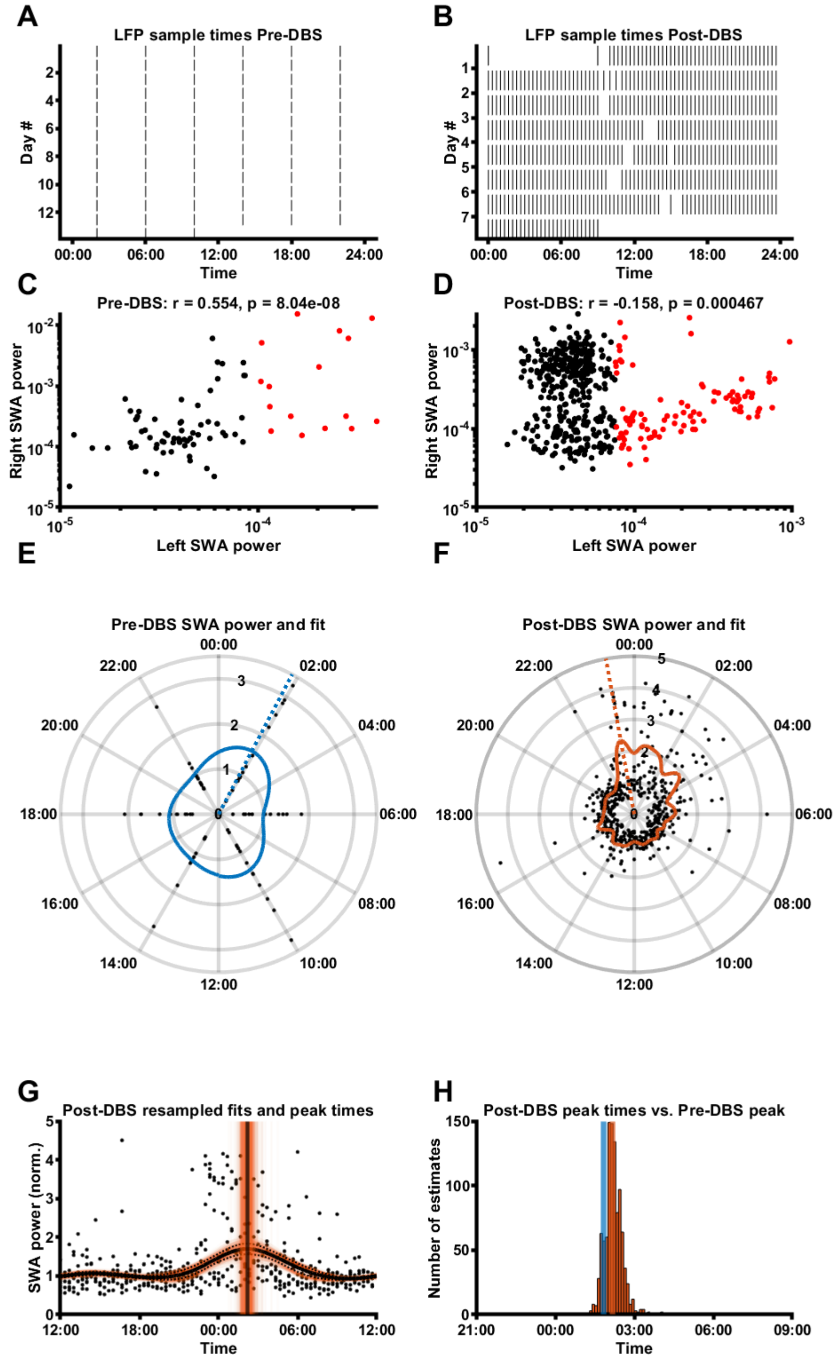

**Figure S2: P001 SWA data overview**

**A, B:** SCC LFP sample times in the Pre-DBS (A) and Post-DBS (B) phase. **C, D:** Correlation of left and right SCC SWA power Pre-DBS (C) and Post-DBS (D). **E, F:** SWA power (normalised to median) of all LFPs for this participant (mean across valid hemispheres), plotted around the 24h diurnal cycle. The coloured line represents a smoothing spline fit to the data; the dashed line represents the time of the maximum night-time (18:00-10:00) peak of the SWA fit. **G:** SWA power (normalised to median) of all LFPs for this participant (mean across valid hemispheres), with a mean±SD fit line (solid black line and dashed black lines) superimposed on 1000 fit lines (thin, orange) obtained through random re-sampling of Post-DBS data according to the sample times Pre-DBS (see Methods for details). Also indicated is the median fit peak estimate (black vertical line) superimposed on 1000 fit peak estimate lines (thin orange vertical lines). **H:** Distribution of fit peak time estimates obtained through the 1000 random resamples of the Post-DBS phase, with the median peak time estimate indicated with the orange vertical line and the fit peak time from Phase B indicated with the blue vertical line.

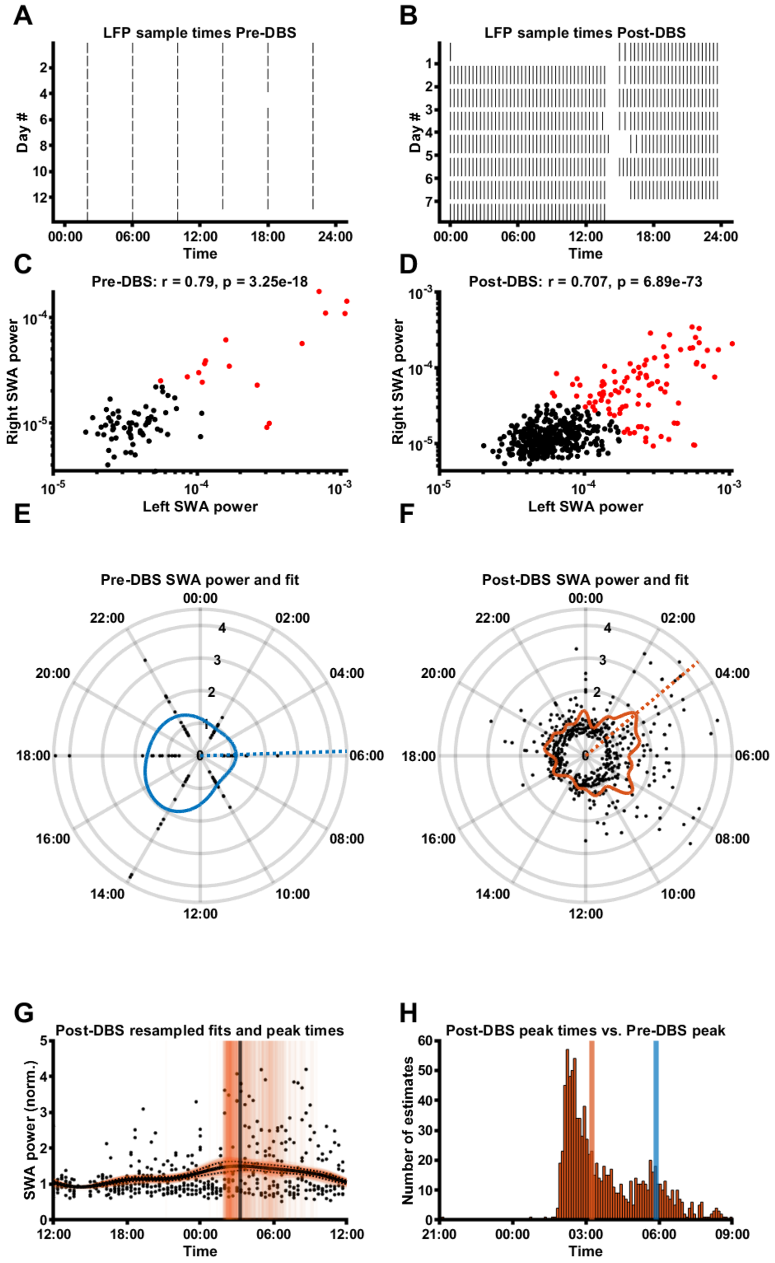

**Figure S3: P002 SWA data overview**

**A, B:** SCC LFP sample times in the Pre-DBS (A) and Post-DBS (B) phase. **C, D:** Correlation of left and right SCC SWA power Pre-DBS (C) and Post-DBS (D). **E, F:** SWA power (normalised to median) of all LFPs for this participant (mean across valid hemispheres), plotted around the 24h diurnal cycle. The coloured line represents a smoothing spline fit to the data; the dashed line represents the time of the maximum night-time (18:00-10:00) peak of the SWA fit. **G:** SWA power (normalised to median) of all LFPs for this participant (mean across valid hemispheres), with a mean±SD fit line (solid black line and dashed black lines) superimposed on 1000 fit lines (thin, orange) obtained through random re-sampling of Post-DBS data according to the sample times Pre-DBS (see Methods for details). Also indicated is the median fit peak estimate (black vertical line) superimposed on 1000 fit peak estimate lines (thin orange vertical lines). **H:** Distribution of fit peak time estimates obtained through the 1000 random resamples of the Post-DBS phase, with the median peak time estimate indicated with the orange vertical line and the fit peak time from Phase B indicated with the blue vertical line.

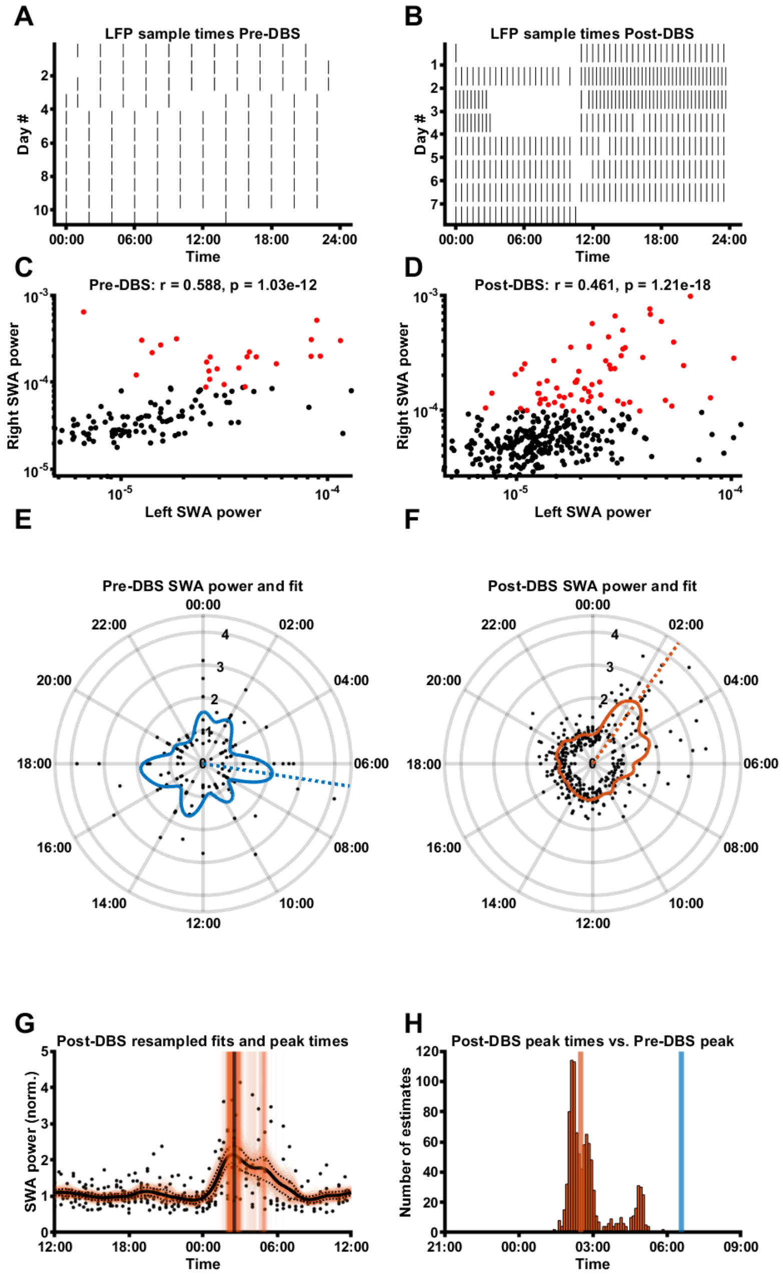

**Figure S4: P003 SWA data overview**

**A, B:** SCC LFP sample times in the Pre-DBS (A) and Post-DBS (B) phase. **C, D:** Correlation of left and right SCC SWA power Pre-DBS (C) and Post-DBS (D). **E, F:** SWA power (normalised to median) of all LFPs for this participant (mean across valid hemispheres), plotted around the 24h diurnal cycle. The coloured line represents a smoothing spline fit to the data; the dashed line represents the time of the maximum night-time (18:00-10:00) peak of the SWA fit. **G:** SWA power (normalised to median) of all LFPs for this participant (mean across valid hemispheres), with a mean±SD fit line (solid black line and dashed black lines) superimposed on 1000 fit lines (thin, orange) obtained through random re-sampling of Post-DBS data according to the sample times Pre-DBS (see Methods for details). Also indicated is the median fit peak estimate (black vertical line) superimposed on 1000 fit peak estimate lines (thin orange vertical lines). **H:** Distribution of fit peak time estimates obtained through the 1000 random resamples of the Post-DBS phase, with the median peak time estimate indicated with the orange vertical line and the fit peak time from Phase B indicated with the blue vertical line.

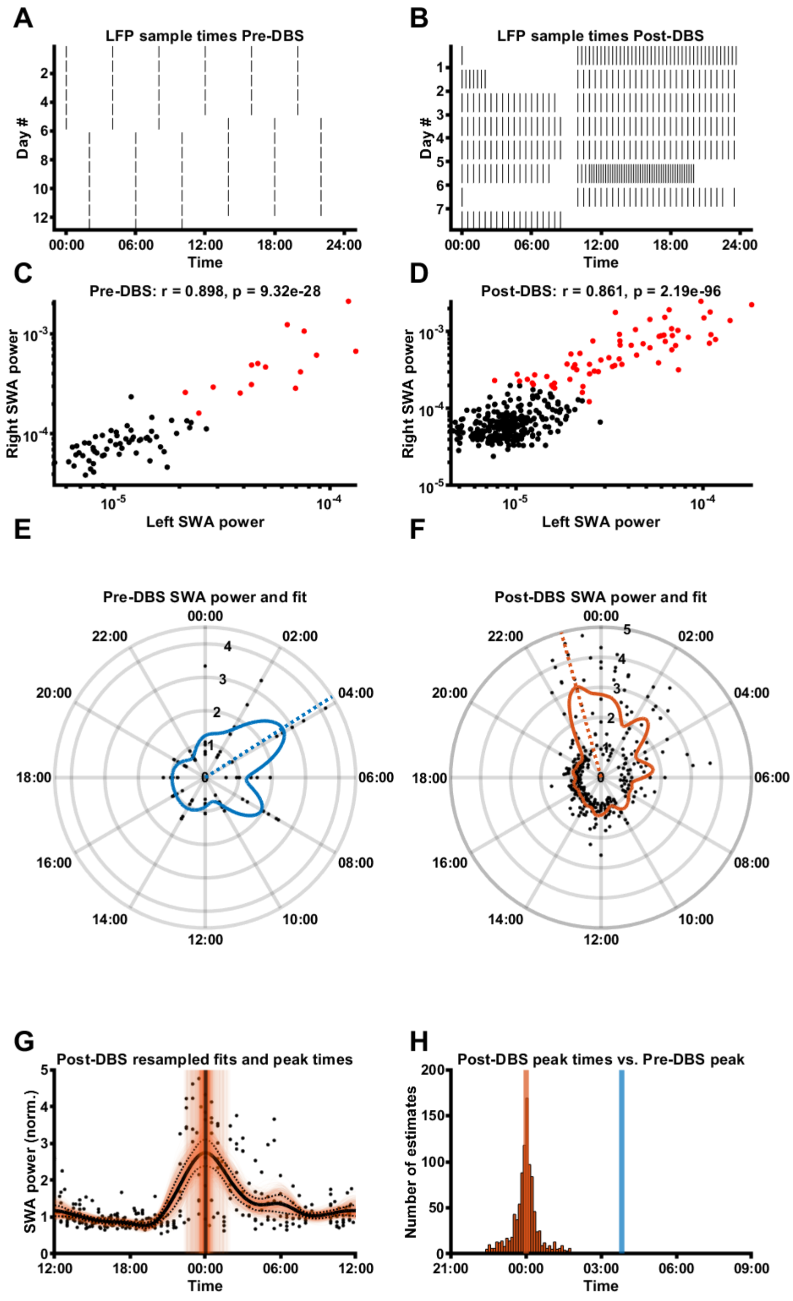

**Figure S5: P004 SWA data overview**

**A, B:** SCC LFP sample times in the Pre-DBS (A) and Post-DBS (B) phase. **C, D:** Correlation of left and right SCC SWA power Pre-DBS (C) and Post-DBS (D). **E, F:** SWA power (normalised to median) of all LFPs for this participant (mean across valid hemispheres), plotted around the 24h diurnal cycle. The coloured line represents a smoothing spline fit to the data; the dashed line represents the time of the maximum night-time (18:00-10:00) peak of the SWA fit. **G:** SWA power (normalised to median) of all LFPs for this participant (mean across valid hemispheres), with a mean±SD fit line (solid black line and dashed black lines) superimposed on 1000 fit lines (thin, orange) obtained through random re-sampling of Post-DBS data according to the sample times Pre-DBS (see Methods for details). Also indicated is the median fit peak estimate (black vertical line) superimposed on 1000 fit peak estimate lines (thin orange vertical lines). **H:** Distribution of fit peak time estimates obtained through the 1000 random resamples of the Post-DBS phase, with the median peak time estimate indicated with the orange vertical line and the fit peak time from Phase B indicated with the blue vertical line.

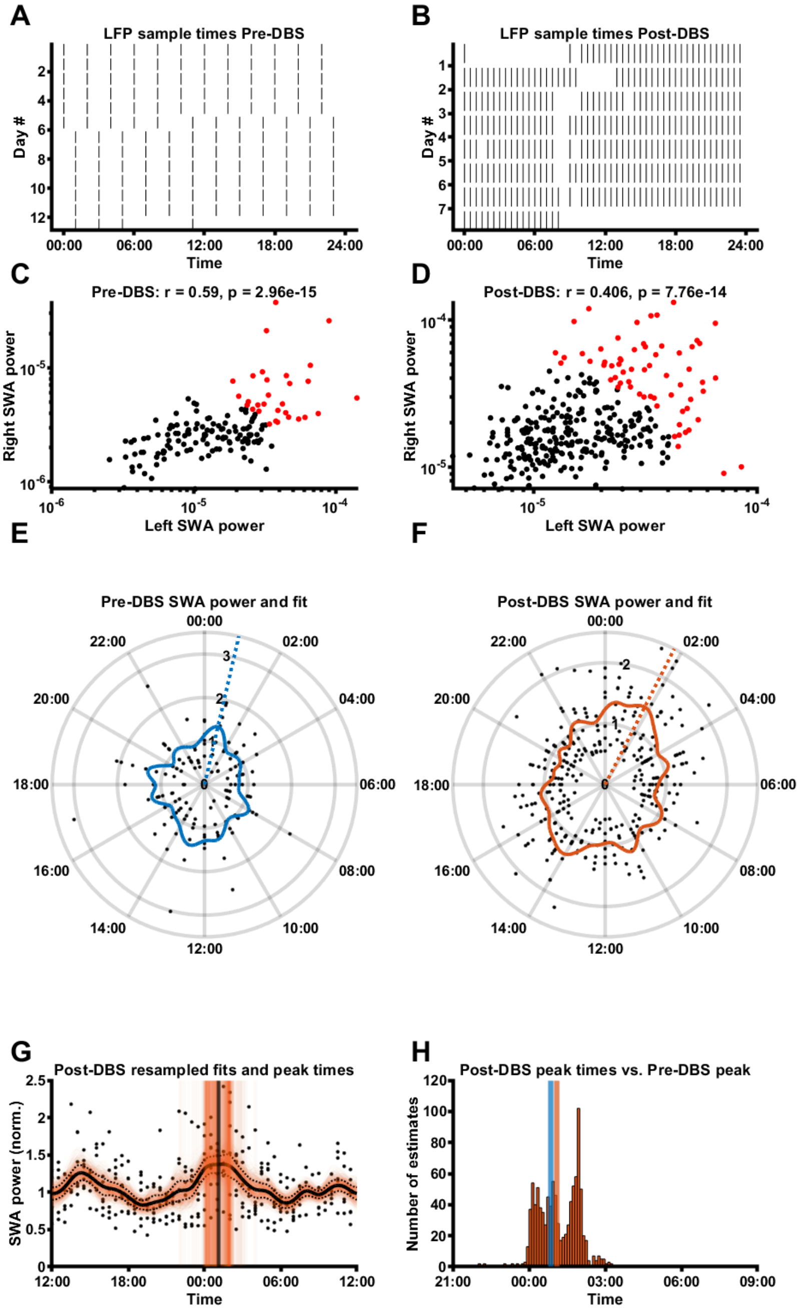

**Figure S6: P005 SWA data overview**

**A, B:** SCC LFP sample times in the Pre-DBS (A) and Post-DBS (B) phase. **C, D:** Correlation of left and right SCC SWA power Pre-DBS (C) and Post-DBS (D). **E, F:** SWA power (normalised to median) of all LFPs for this participant (mean across valid hemispheres), plotted around the 24h diurnal cycle. The coloured line represents a smoothing spline fit to the data; the dashed line represents the time of the maximum night-time (18:00-10:00) peak of the SWA fit. **G:** SWA power (normalised to median) of all LFPs for this participant (mean across valid hemispheres), with a mean±SD fit line (solid black line and dashed black lines) superimposed on 1000 fit lines (thin, orange) obtained through random re-sampling of Post-DBS data according to the sample times Pre-DBS (see Methods for details). Also indicated is the median fit peak estimate (black vertical line) superimposed on 1000 fit peak estimate lines (thin orange vertical lines). **H:** Distribution of fit peak time estimates obtained through the 1000 random resamples of the Post-DBS phase, with the median peak time estimate indicated with the orange vertical line and the fit peak time from Phase B indicated with the blue vertical line.

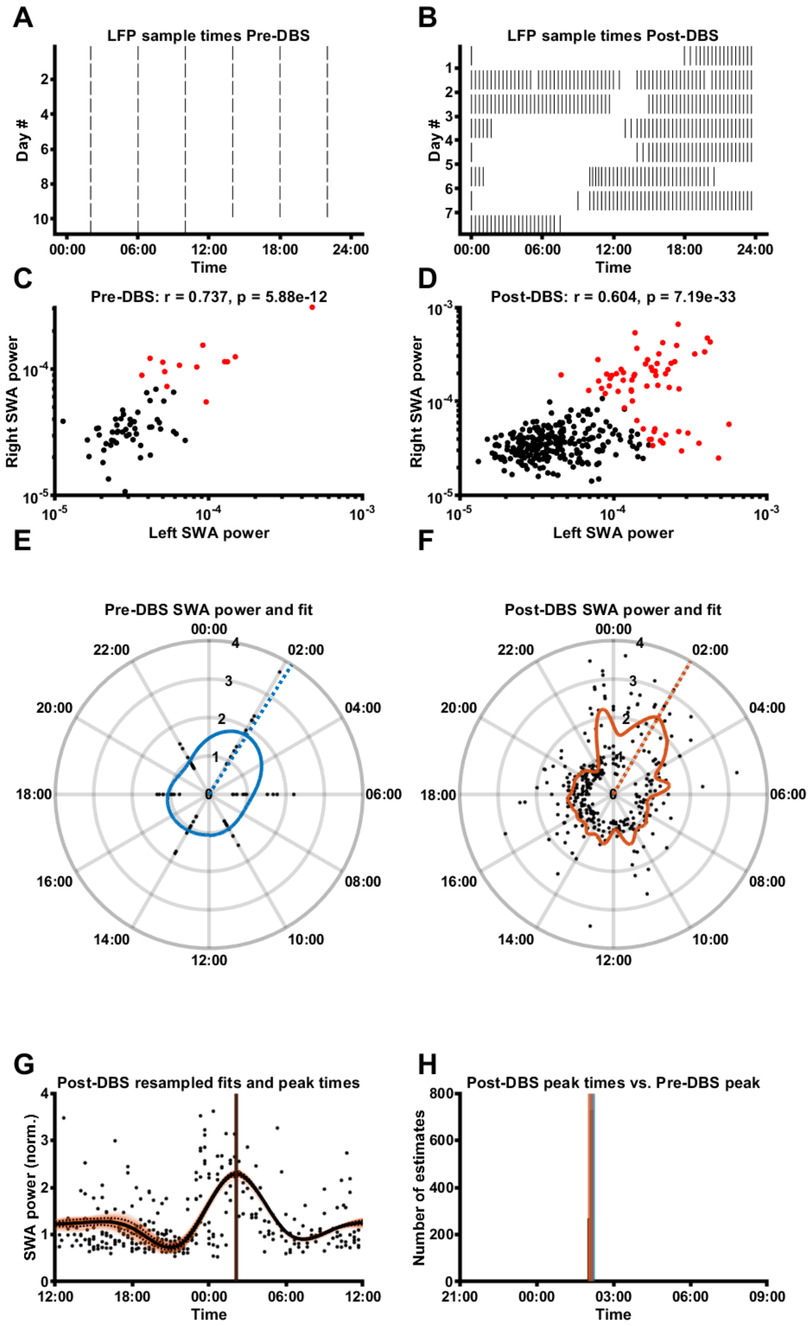

**Figure S7: P006 SWA data overview**

**A, B:** SCC LFP sample times in the Pre-DBS (A) and Post-DBS (B) phase. **C, D:** Correlation of left and right SCC SWA power Pre-DBS (C) and Post-DBS (D). **E, F:** SWA power (normalised to median) of all LFPs for this participant (mean across valid hemispheres), plotted around the 24h diurnal cycle. The coloured line represents a smoothing spline fit to the data; the dashed line represents the time of the maximum night-time (18:00-10:00) peak of the SWA fit. **G:** SWA power (normalised to median) of all LFPs for this participant (mean across valid hemispheres), with a mean±SD fit line (solid black line and dashed black lines) superimposed on 1000 fit lines (thin, orange) obtained through random re-sampling of Post-DBS data according to the sample times Pre-DBS (see Methods for details). Also indicated is the median fit peak estimate (black vertical line) superimposed on 1000 fit peak estimate lines (thin orange vertical lines). **H:** Distribution of fit peak time estimates obtained through the 1000 random resamples of the Post-DBS phase, with the median peak time estimate indicated with the orange vertical line and the fit peak time from Phase B indicated with the blue vertical line.

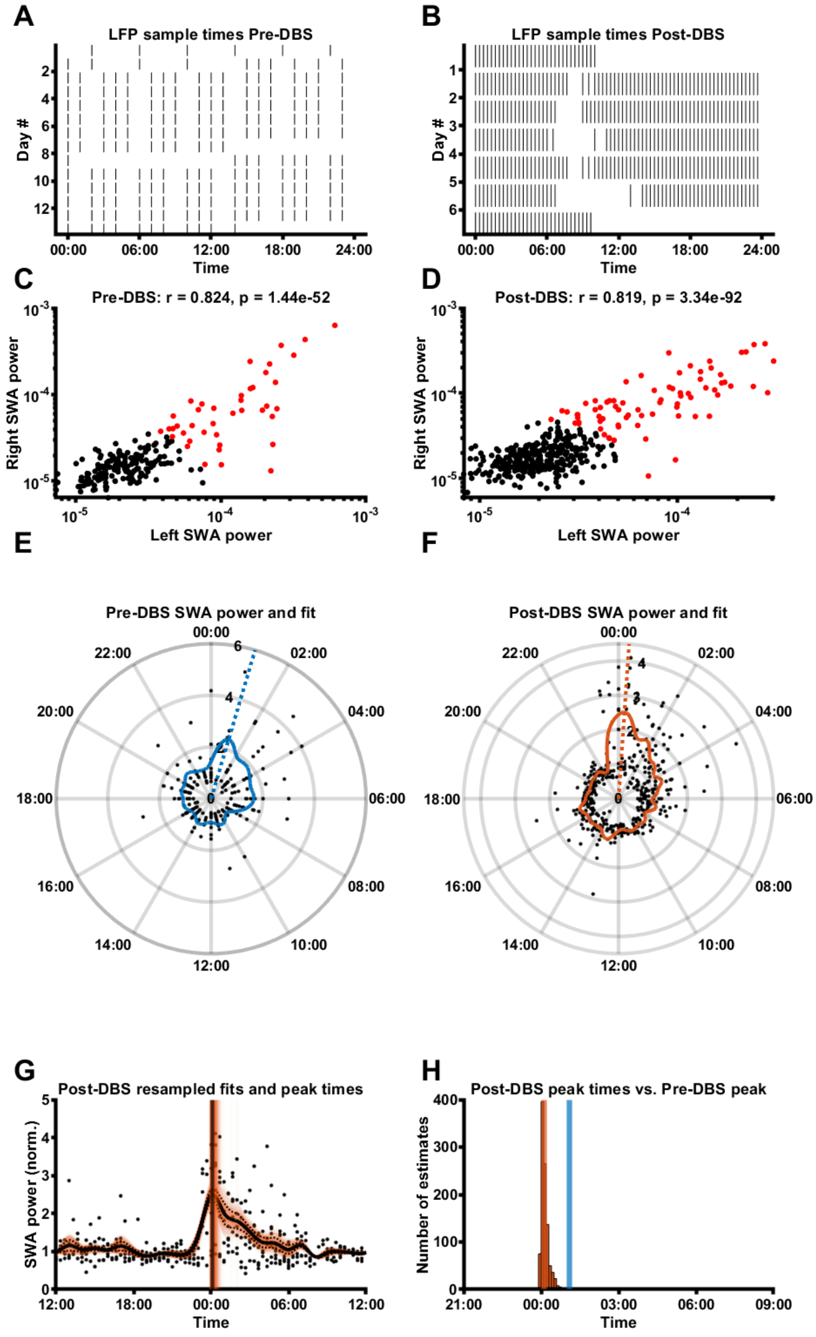

**Figure S8: P007 SWA data overview**

**A, B:** SCC LFP sample times in the Pre-DBS (A) and Post-DBS (B) phase. **C, D:** Correlation of left and right SCC SWA power Pre-DBS (C) and Post-DBS (D). **E, F:** SWA power (normalised to median) of all LFPs for this participant (mean across valid hemispheres), plotted around the 24h diurnal cycle. The coloured line represents a smoothing spline fit to the data; the dashed line represents the time of the maximum night-time (18:00-10:00) peak of the SWA fit. **G:** SWA power (normalised to median) of all LFPs for this participant (mean across valid hemispheres), with a mean±SD fit line (solid black line and dashed black lines) superimposed on 1000 fit lines (thin, orange) obtained through random re-sampling of Post-DBS data according to the sample times Pre-DBS (see Methods for details). Also indicated is the median fit peak estimate (black vertical line) superimposed on 1000 fit peak estimate lines (thin orange vertical lines). **H:** Distribution of fit peak time estimates obtained through the 1000 random resamples of the Post-DBS phase, with the median peak time estimate indicated with the orange vertical line and the fit peak time from Phase B indicated with the blue vertical line.

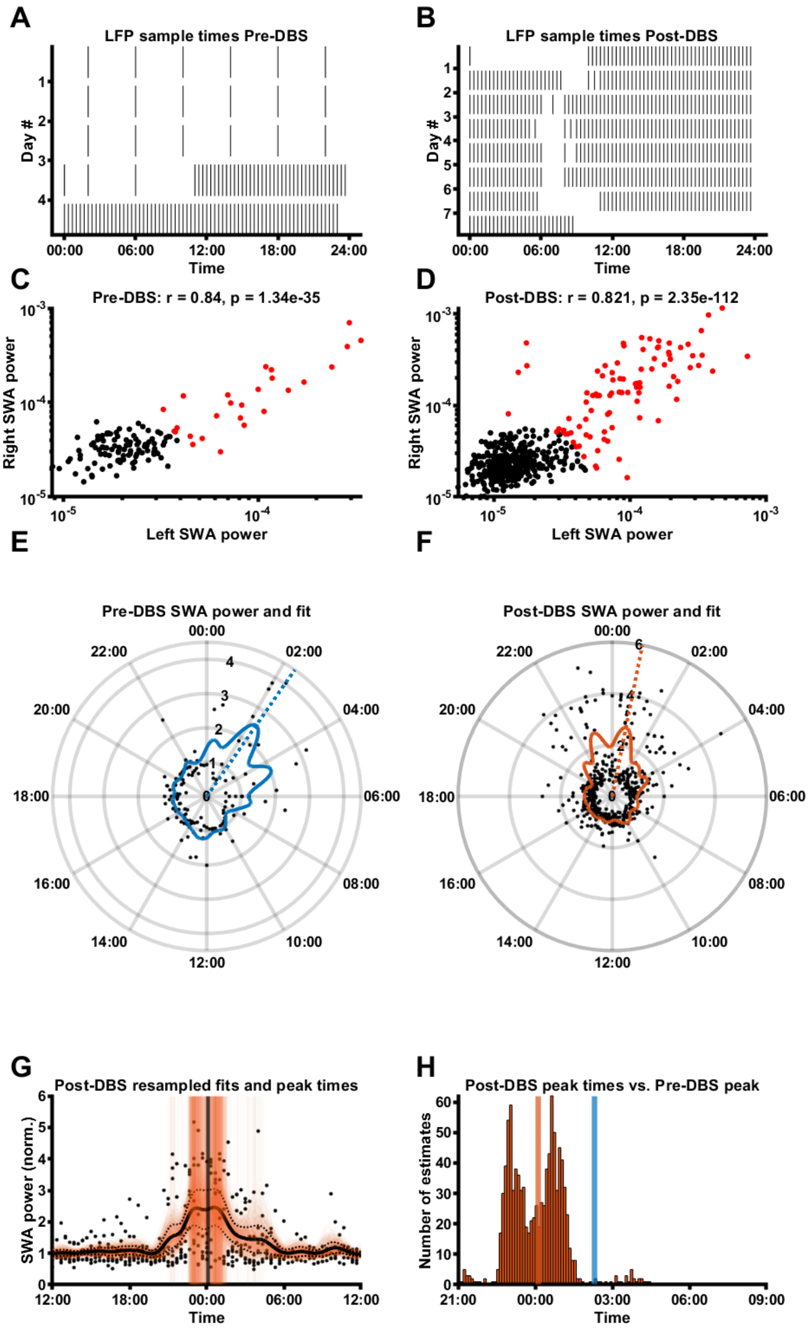

**Figure S9: P008 SWA data overview**

**A, B:** SCC LFP sample times in the Pre-DBS (A) and Post-DBS (B) phase. **C, D:** Correlation of left and right SCC SWA power Pre-DBS (C) and Post-DBS (D). **E, F:** SWA power (normalised to median) of all LFPs for this participant (mean across valid hemispheres), plotted around the 24h diurnal cycle. The coloured line represents a smoothing spline fit to the data; the dashed line represents the time of the maximum night-time (18:00-10:00) peak of the SWA fit. **G:** SWA power (normalised to median) of all LFPs for this participant (mean across valid hemispheres), with a mean±SD fit line (solid black line and dashed black lines) superimposed on 1000 fit lines (thin, orange) obtained through random re-sampling of Post-DBS data according to the sample times Pre-DBS (see Methods for details). Also indicated is the median fit peak estimate (black vertical line) superimposed on 1000 fit peak estimate lines (thin orange vertical lines). **H:** Distribution of fit peak time estimates obtained through the 1000 random resamples of the Post-DBS phase, with the median peak time estimate indicated with the orange vertical line and the fit peak time from Phase B indicated with the blue vertical line.

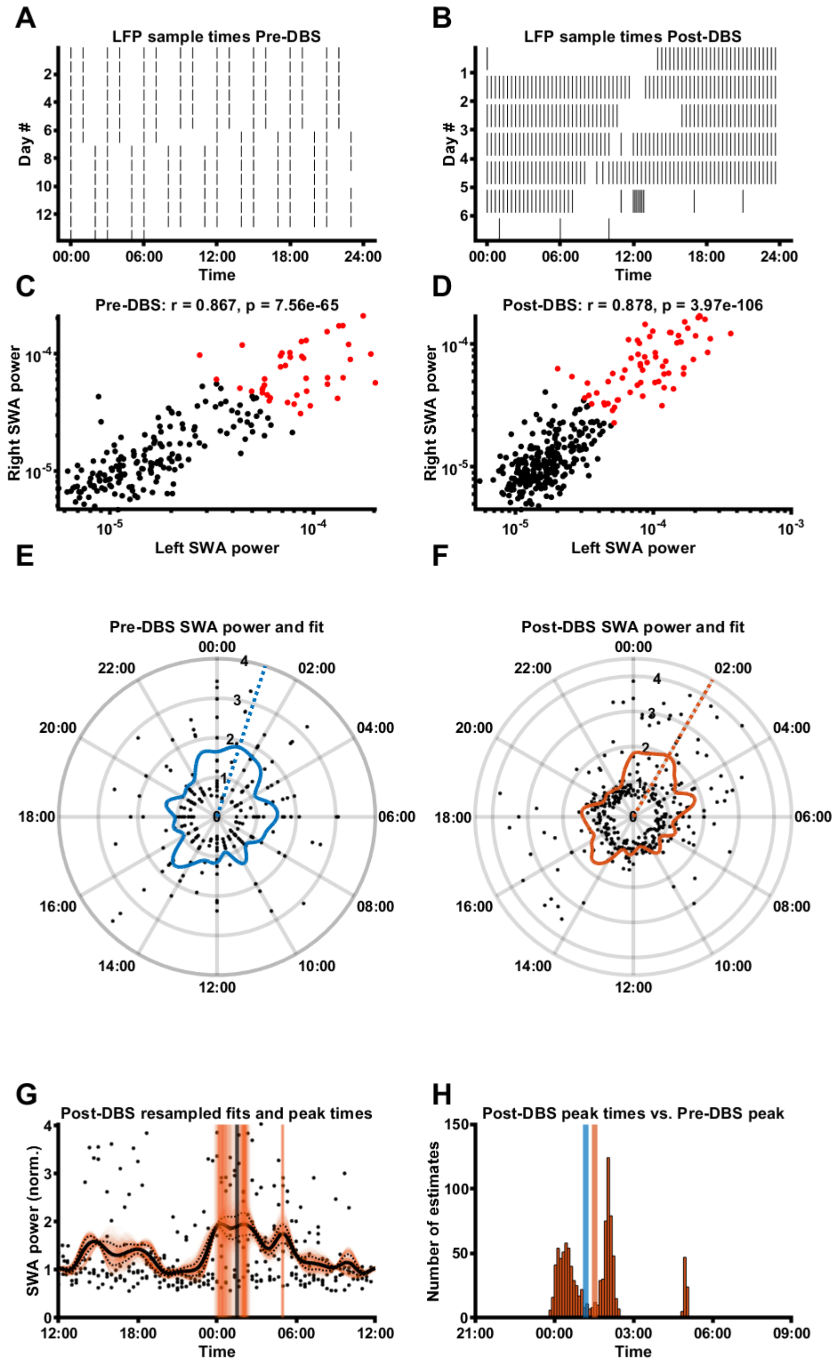

**Figure S10: P009 SWA data overview**

**A, B:** SCC LFP sample times in the Pre-DBS (A) and Post-DBS (B) phase. **C, D:** Correlation of left and right SCC SWA power Pre-DBS (C) and Post-DBS (D). **E, F:** SWA power (normalised to median) of all LFPs for this participant (mean across valid hemispheres), plotted around the 24h diurnal cycle. The coloured line represents a smoothing spline fit to the data; the dashed line represents the time of the maximum night-time (18:00-10:00) peak of the SWA fit. **G:** SWA power (normalised to median) of all LFPs for this participant (mean across valid hemispheres), with a mean±SD fit line (solid black line and dashed black lines) superimposed on 1000 fit lines (thin, orange) obtained through random re-sampling of Post-DBS data according to the sample times Pre-DBS (see Methods for details). Also indicated is the median fit peak estimate (black vertical line) superimposed on 1000 fit peak estimate lines (thin orange vertical lines). **H:** Distribution of fit peak time estimates obtained through the 1000 random resamples of the Post-DBS phase, with the median peak time estimate indicated with the orange vertical line and the fit peak time from Phase B indicated with the blue vertical line.

### Supplementary tables

**Supplementary Table 1: Patient medication intake schedule (kept constant between study phases)**

| Patient ID | Medication / supplement | Regime |
|---|---|---|
| **P001** | Bupropion extended release 300 mg | qDay |
| | Bupropion 75 mg | qDay |
| | Bupropion extended release 150 mg | q24hr |
| | Cephalexin 500 mg | QID |
| | Cholecalciferol 2,000 IU | qDay |
| | Clonazepam 0.5 mg | qHS, PRN |
| | Ferrous gluconate 325 mg | qDay |
| | l-methylfolate15 mg | qDay |
| **P002** | Citalopram | |
| | Desvenlafaxine 50 mg extended release | qDay |
| | Docusate 100 mg | BID, PRN |
| | Lamotrigine 200 mg | |
| | Ondansetron 4 mg | q8hr, PRN |
| **P003** | Clonazepam 2 mg | qHS |
| | Dextromethorphan 30 mg | qDay |
| | Diazepam 5 mg | q8hr, PRN |
| | Diltiazem 180 mg | qDay |
| | Diphenhydramine 25 mg | qHS |
| | Docusate 100 mg | q12hr |
| | Escitalopram 40 mg | qDay |
| | Melatonin 5 mg | qHS |
| | Omega-3 polyunsaturated fatty acids 1,000 mg | TID |
| | Ondansetron 4 mg | q8hr, PRN |
| | Acetaminophen/aspirin/caffeine (Excedrin) 2 tab(s) | q6hr |
| **P004** | Lunesta 3mg | qHS,PRN |
| | Gabapentin 300 mg | TID |
| | l-methylfolate | qDay |
| | Fetzima 120 mg, | qDay |
| | Ativan 1 mg, | BID, PRN |
| | Quetiapine 200 mg | qPM |
| | Tramadol 50 mg | q4hr |
| **P005** | Clonazepam 1.5 mg | qHS |
| | Levothyroxine 175 mcg | qDay |
| | Nicotine 1 patch(es), Topical | q24hr |
| | Nortriptyline 150 mg | qHS |
| | Quetiapine 300 mg | qHS |

| | | |
|---|---|---|
| | Sertraline 50 mg | qDay |
| | Sertraline 100 mg | qDay |
| **P006** | Amitriptyline 350 mg | |
| | Lorazepam 1 mg | TID, PRN |
| **P007** | Adderall 30mg | |
| | Clonazepam 1 mg | |
| | Pristiq | qDay |
| **P008** | Bupropion 450 mg | q24hr |
| | Lunesta 1 mg, | qHS, PRN |
| | Lorazepam 0.5 mg | BID, PRN |
| | Trokendi XR 200 mg | qDay |
| **P009** | Klonopin 0.5 mg | BID |
| | Cymbalta 120 mg | qDay |
| | Lithium 300 mg | |

**Supplementary Table 2: Pearson's correlation coefficients of Pre-DBS Hamilton scores and sleep subscales vs. SWA fit peak time and height. Bonferroni-corrected alpha level = 0.01.**

| | **SWA peak time** | | **SWA peak height** | |
|---|---|---|---|---|
| | ***r*** | ***p* (α=0.01)** | ***r*** | ***p* (α=0.01)** |
| **Hamilton score** | -0.567 | 0.143 | 0.418 | 0.302 |
| **Initial insomnia** | -0.439 | 0.276 | -0.027 | 0.950 |
| **Midnight insomnia** | -0.310 | 0.455 | 0.278 | 0.506 |
| **Morning insomnia** | -0.501 | 0.206 | -0.074 | 0.862 |
| **Hypersomnia** | -0.132 | 0.755 | 0.341 | 0.408 |

**Supplementary Table 3: Pearson's correlation coefficients of the Pre- vs. Post-DBS difference in Hamilton scores and sleep subscales vs. the Pre- vs. Post-DBS difference in SWA fit peak time and height. Bonferroni-corrected alpha level = 0.01.**

| | **Δ SWA peak time** | | **Δ SWA peak height** | |
|---|---|---|---|---|
| | ***r*** | ***p* (α=0.01)** | ***r*** | ***p* (α=0.01)** |
| **Δ Hamilton score** | 0.004 | 0.993 | 0.130 | 0.759 |
| **Δ Initial insomnia** | -0.204 | 0.628 | -0.061 | 0.885 |
| **Δ Midnight insomnia** | 0.022 | 0.958 | -0.359 | 0.382 |
| **Δ Morning insomnia** | -0.636 | 0.090 | -0.071 | 0.867 |
| **Δ Hypersomnia** | -0.158 | 0.709 | 0.712 | 0.048 |

**Supplementary Table 4: Pearson's correlation coefficients of the Pre- vs. Post-DBS difference in Hamilton scores and sleep subscales vs. the Pre- vs. Post-DBS difference in spindle density and amplitude. Bonferroni-corrected alpha level = 0.01.**

| | **Δ Spindle density** | | **Δ Spindle amplitude** | |
|---|---|---|---|---|
| | ***r*** | ***p* (α=0.01)** | ***r*** | ***p* (α=0.01)** |
| **Δ Hamilton score** | 0.231 | 0.583 | 0.236 | 0.574 |
| **Δ Initial insomnia** | -0.579 | 0.133 | 0.188 | 0.656 |
| **Δ Midnight insomnia** | 0.481 | 0.228 | 0.117 | 0.783 |
| **Δ Morning insomnia** | -0.152 | 0.719 | -0.458 | 0.254 |
| **Δ Hypersomnia** | 0.498 | 0.209 | 0.214 | 0.611 |
